# Modulating Factors Affecting Sports-Related Concussion Exposures: A Systematic Review and Analysis

**DOI:** 10.1101/2023.03.08.23286974

**Authors:** Rachel Edelstein, John Darrell Van Horn

## Abstract

In the United States, the Centers for Disease Control and Prevention estimates that 1.6-to-3.8 million concussions occur annually in sports. This quantitative meta-analysis of one hundred and twenty-one sports-related concussion studies, across fourteen youth sports, investigated the contributions of statistical constructs, and diagnostic methods, considering the impact of nationwide laws and education, as well as distinguished injury surveillance techniques, in this widely diverse literature. Concussion study research designs were found to have differing diagnostic, statistical, and methodological techniques. Among less frequently studied, non-high-contact, sports, a meta-analysis investigated relative rates of concussion and sex differences. Findings indicated considerable variation in reported concussion incidence rates due to the sport played, but also due to the number of study authors listed and the proportion of female athletes represented. Such factors likely influence the reported incidence of sports-related concussions, especially in sports not typically associated with a high risk of head injuries. To better capture the incidence of sports-related head injuries and concussion, a universal data repository for youth concussions might be established to provide an ongoing, comprehensive, and accurate picture of concussions in youth and collegiate athletics.

## INTRODUCTION

In the United States, the Centers for Disease Control and Prevention (CDC) estimates that annually, 1.6-to-3.8 million sports-related concussions (SRC) occur in sports. This may likely be a large underestimation due to poor and inconsistent reporting within as well as across various sporting domains.^1^ In fact, for those between the ages 15-to-24, sports are ranked behind motor vehicle collisions as the leading cause of traumatic brain injury.^2^ In contrast to professional sports, the focus in the media and in research on SRC has rarely concerned high-school and collegiate athletes,(3, 4) in spite there being nearly 8 million participants in youth sports annually.^5^ Many youth sporting activities are now starting at younger ages than twenty years ago, and 60% of boys and 47% of girls are on a team by the age of six. Whereas only 20% of youth athletes continue to play after fifteen years of age, in those where records exist, evidence suggests injuries experienced during youth sports may potentiate the development of neurodegenerative diseases in later adulthood.^6^

Consistent and effective protocols concerning suspected SRC in youth sports have been irregularly investigated. In 2010, the pioneering “Zackery Lystedt Law”^1^ was proposed in the State of Washington, requiring that young athletes who are suspected of suffering a SRC or serious head contact must be removed from play immediately. Additionally, athletes are only permitted to return with written medical consent from a licensed physician or healthcare provider.^7^ This legislation came about in response to the growing participation in youth sports and the increasing awareness of how devastating head injuries can be. By 2014, 48 US states had SRC education training mandates whereas, by 2017, all 50 states required SRC awareness education by coaching staffs. Nevertheless, for many requirements its it unclear concerning who should receive such instruction and how frequently it should be re-administered.^8^ Currently, the only sports-related concussion-education requirement for coaches a course designed by the CDC that requires renewal every three years, and though coaches reportedly felt more confident in identifying these injuries after these educational courses, it cannot be comparable to the presence of a medical professional.

Consequently, although the number of athletes who participate in youth compared to collegiate sports is three times as great, there is no official injury surveillance for recreational or competitive youth sports. Youth athletes, specifically those who play in a community-based league such as a “travel” or “club” league, are not required to have an athletic trainer on staff, leaving the coach and athlete responsible for reporting injuries.^11^ In a 2018 study, a sample of N=845 youth ages 12–17 years responded to the web-based Youth Styles survey, which sought to understand whether athletes pursued care after a SRC. Their findings indicated that a physician never evaluated 30% of athletes’ post-injury and many of these injuries were sustained in a community-based sports setting (∼36%).^11^

A standard statistical scaling approach conducted when evaluating SRC exposure risk is through Athletic Exposure (AE) which gauges the risk of injury to the athlete based on a particular measurement of time. Specifically, AE calibrates the risk of sustaining an injury over a specific period (e.g., 1 season or 1,000 hours) and is viewed as a more intuitive measure of injury incidence for non-scientists.^12^ Although this metric is relatively new in investigating SRC injury risk, generalizing athletic exposures can have limitations and sometimes affect the results in how those sports are viewed. First, many researchers isolate sports with higher AE incidence rates, therefore disregarding SRC in other sports due to their low incidence rate. In high performance athletes it’s not uncommon to believe that 1,000 hours could be reached within months. For example, in a sport such as tennis with a relatively low incidence rate (e.g., 0.05-0.15 AE by 1000 hrs) it can be assumed a SRC within this sport is less likely, enforcing the idea that a SRC can’t truly happen in other non-contact sports.

A second disadvantage with this metric, while useful, may tend to differ slightly from one researcher to the next due to variation in the underlying baseline considered. For instance, in youth and college sports, not every athlete receives the same amount of playing time, thus, using this approach can only apply to those athletes who compete in every game and practice. A further concern with this method can be that researchers differ in when “the clock starts”, meaning, some researchers consider the AE hours to begin when an athlete enters competition and practice. In contrast, other researchers consider the clock to start the moment an athlete leaves the training room – including non-competition time, practice time, competition time, as well as on- and off-field warm-ups.^12^ These differences in approaches can be one explanation for the inconsistent conclusions concerning SRC in the literature as well as the widely varying policies within and between sports.

In reviewing the SRC literature, many published meta-analytic review studies suffered uncertain validity and poor reliability due to several experimental design and statistical insufficiencies that have led to statistical validity generalization concerns. As seen in Table 1, most studies had fewer than 20 studies included in the final quantitative review. Similar trends were notes by other authors noticed specific factors to cause these limitations such as, unclear reporting of measurement and data collection protocols, cohort demographics, and unreported confounding external variables. Moreover, the situation-by-treatment interactions (e.g., treatment conditions, time, AE hours, location, choice of concussion assessment, statistical protocols, treatment administration, investigator, timing, scope and extent of measurement), together with how those AE sample risk estimates may have certain features that interact with the independent variable contribute to study potentially limit clinical and statistical generalizability.

**Table 1.**
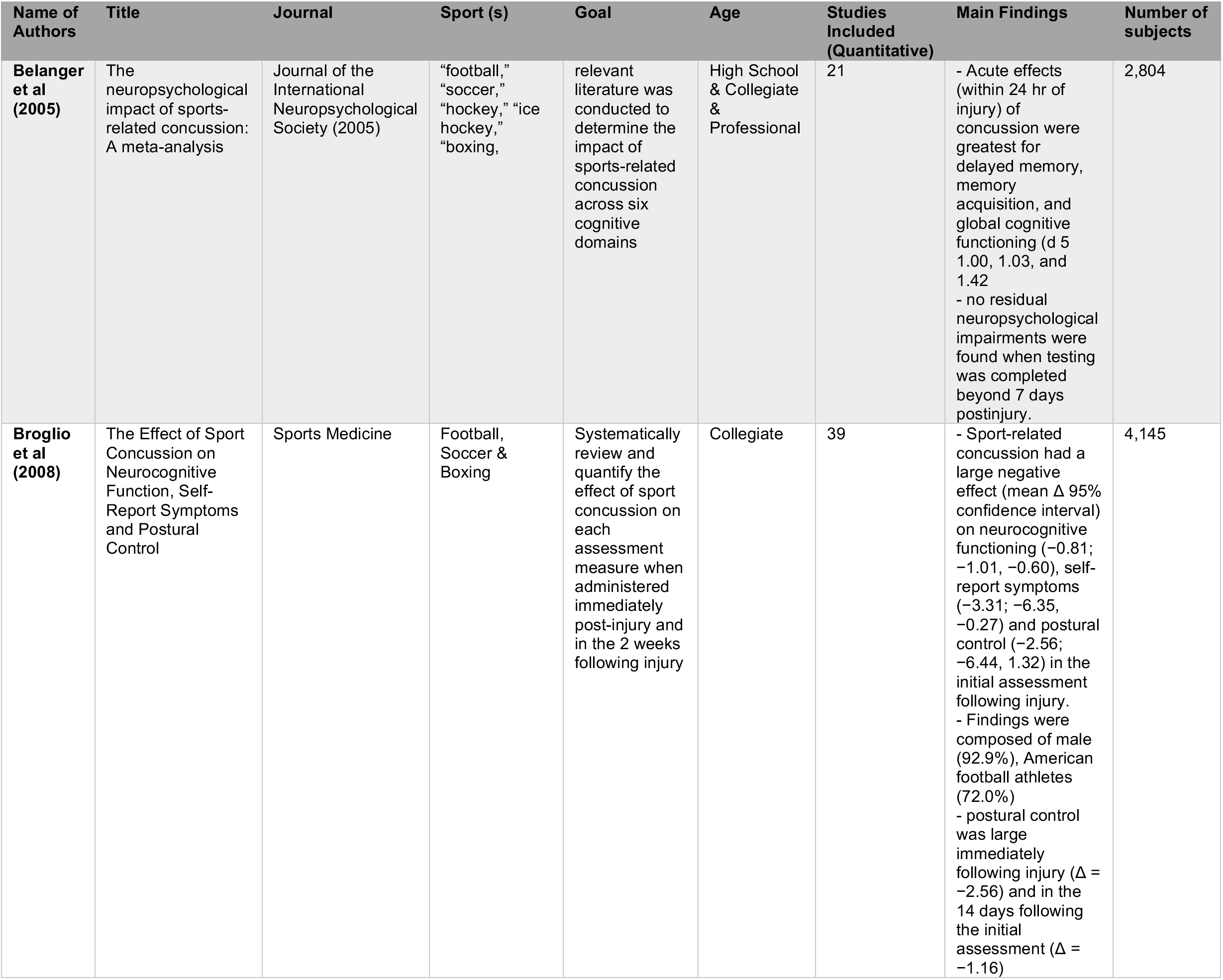
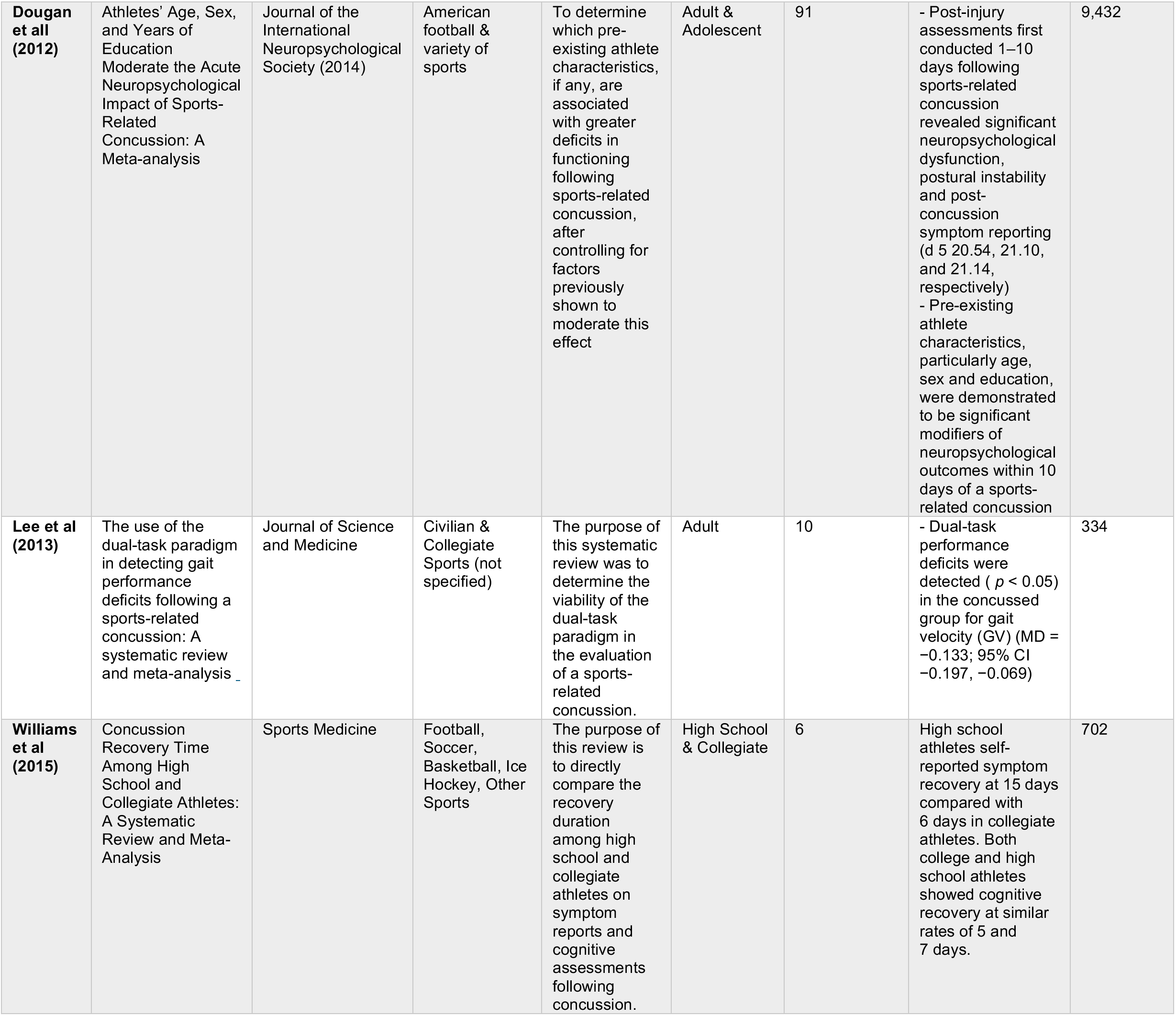
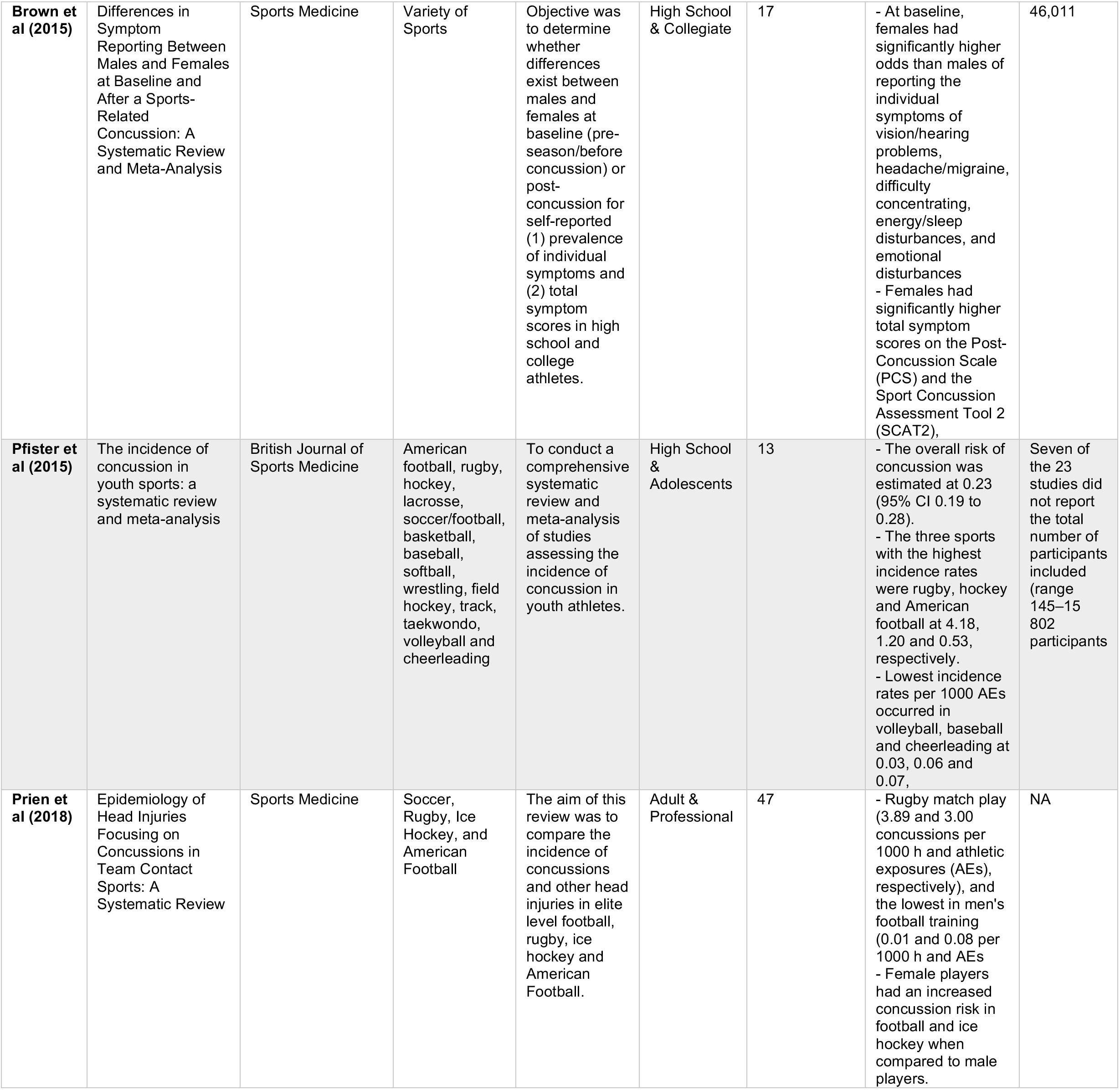
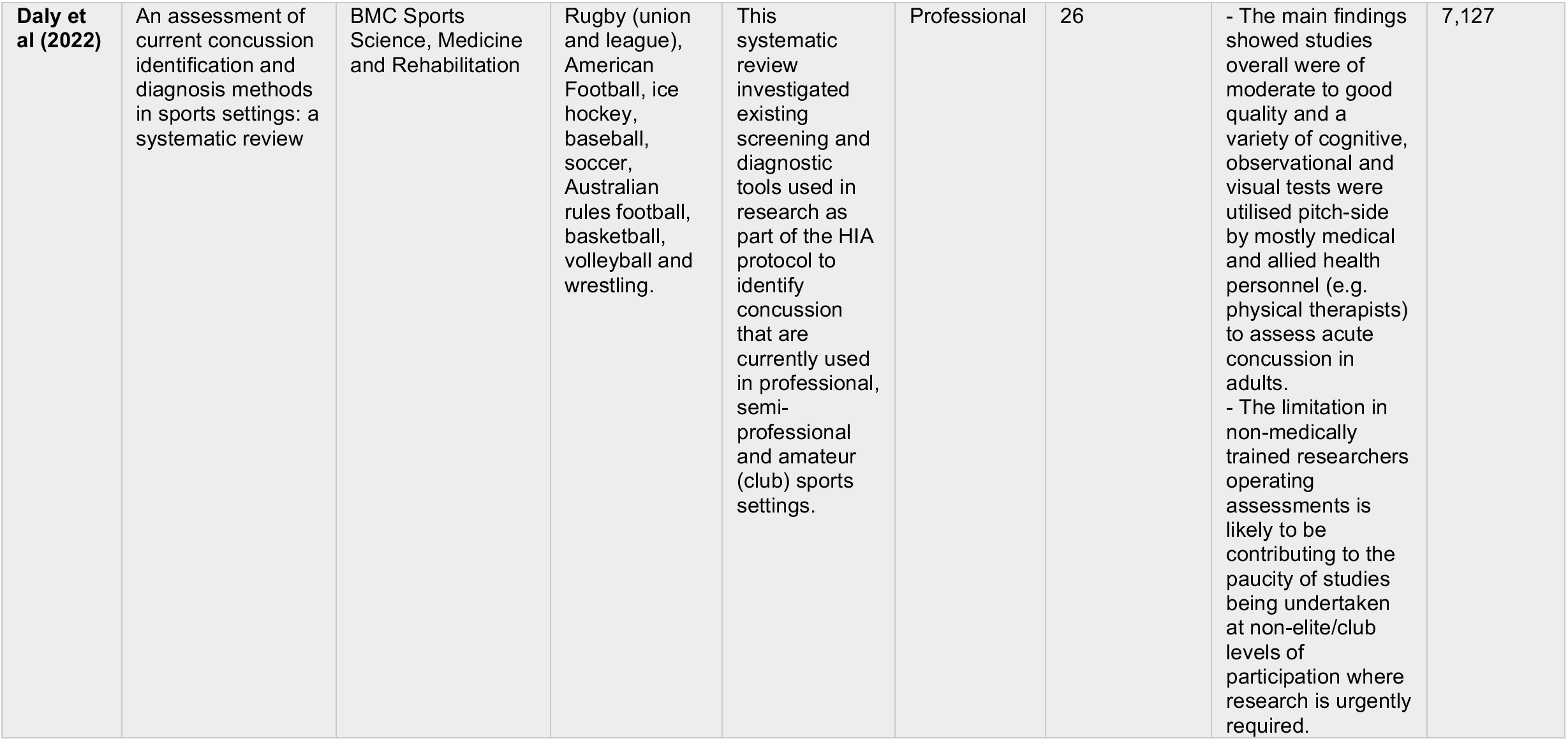
Past Meta-Analytic Reviews of Sports Concussion

## LITERATURE REVIEW

A literature review is presented here involving the range of low or non-contact, sex-comparable sports (Baseball, Cricket, Softball, Basketball, Cheerleading, Gymnastics, Crew/Rowing, Cross Country, Track and Field, Diving, Swimming, Equestrian, Field Hockey, Lacrosse, Surfing, Snowboarding, Skiing, Luge, Tennis, Water Polo and Volleyball) to review the representative of many youth or community sports activities in the United States. The purpose of this review was to emphasize different SRC risks, mechanisms and findings within the current literature. It is important to note that a number of these sports have only modest research available, current media reporting’s were mentioned to emphasize the consequences of SRC. As noted above, youth sports were specifically excluded where SRC are particularly well documented (Football, Ice Hockey, Soccer, Wrestling and Rugby), where they a comprehensive collection of research studies in the literature already, and where their inclusion would have tended to greatly overshadow effects present in these other sporting domains.

### Baseball

Limited SRC data exists within youth baseball athletes, let alone the collegiate and/or professional ranks. In a study that compared youth baseball, football, and hockey players, it was observed that sports-related SRC in adolescents is often associated with widespread changes in white matter microstructure integrity up to two months following injury.^13^ Green et al. have noted that a frequent mechanism of SRC injury for baseball players is contact with a batted ball (29.3%)^14^ – in which velocity can reach up to 150 mph once stuck by a batter. Though sports such as baseball have a variety of mechanisms of injury but relatively low-SRC incidence (i.e., in terms of AE), SRC symptoms in baseball were reported as being like high-contact sports such as football or hockey.

### Basketball

As of 2016, basketball is the fastest growing sport by participation in the United States.^15^ Basketball, for either sex, has not traditionally been considered a high SRC risk sport, yet with growing participation and increasing intensity within the athletic community, basketball athletes of both sexes are reporting higher incidence of SRC than even wrestling and soccer.^16^ Specifically, female basketball athletes have reported to have a more substantial risk for SRC. A large-scale study investigating these sex differences reported rates of 7.28 SRC per 1000 AE for boys while an even higher incidence of 8.30 SRC per 1000 AE for females girls.^17^

### Cricket

As in baseball, a cricket ball weighs between 5.5 and 5.75 ounces and can be delivered by international bowlers at speeds over 90 mph/145 kph.^18^ Notably, while famously popular in the United Kingdom and in countries across the former British Empire, cricket in the United States is a sport with upwards of 200,000 players across the country.^20^ Additionally, an assumed exposure rate of SRC, 2.3 and 2.0 SRC per 1,000 player days in elite men’s and women’s cricket, has been reported. ^21^ Consequently, cricket has one of the highest mortalities as a resulting from head injury; with 52 deaths being recorded between 1965 and 2016. ^22^ At the elite level, cricket-related head injuries are typically thought to account for 3–5% of all injuries, but have been reported as high as 25% of all injuries.^23^ Regardless of dangers at the adult level, statistics for exposure risk in youth cricket leagues for any country are rare or non-existent.

### Cheerleading

It is estimated that around 3.8 million children ages 6 and older participated in either cheerleading or cheer-related gymnastics in 2016 in the United States, increasing from 3.2 million in 2007.^24^ While gymnastics (see below) is a sanctioned sport within the National Collegiate Athletics Association (NCAA) in the US, cheerleading is not. This means that the sport has particularly poor reporting for SRC, despite the inclusion of many moves with gymnastic components (e.g., where someone is lifted off the ground, conducts acrobatic maneuvers in the air, and is caught by others). If officially recognized as an NCAA sport, cheerleading would have access to the same benefits as other sports, including resources for better facilities, mandatory SRC education, athletic trainer availability, baseline neurocognitive testing, and inclusion in injury surveillance systems.^25^ On average cheerleaders with SRC were withheld from sport for 26.2 days and where 38% experienced prolonged recovery in excess of 28 days. ^25^ Collegiate cheerleaders appear more likely to sustain a SRC than cheerleaders at the youth level ^26^, although, as noted, SRC reporting in cheerleading is likely unreliable.

### Gymnastics

As in cheerleading, the demand for strength and flexibility is crucial in gymnastics. Furthermore, the recovery from a SRC can be especially challenging due to the nature of returning to positions of inversion. Over the last decade, gymnastic head injuries have been a more frequent topic after a survey of over 300 former NCAA collegiate gymnasts found that 42% reported some type of SRC history, wherein a large number reported seeking mental health treatment post-injury (32% vs. 23%; p = 0.03), and/or having an anxiety disorder diagnosis (16% vs. 9%; p = 0.01). ^29^ Additionally, gymnastics have experienced a steady increase in SRC frequency. From 1998-2004, the average rate of SRC per 1,000 AEs was 0.16.^30^ From 2004-11, that rate for gymnastics specifically increased to 0.7 SRC per 1,000 AEs. ^30^

### Crew/Rowing

Competitive rowing, or “crew”, has historically been a relatively small sport, but has seen considerable growth at the high school-aged level in the United States.^32^ Data on injuries in non-elite high school rowers is lacking and the existing rowing injury epidemiology literature comprises mostly reports from the collegiate, senior, and master’s levels.^33^ While rowing is typically not associated with risk for SRC, head injuries do occur from contact, including with other athletes and equipment (*e.g.,* being accidentally hit with an oar). A retrospective analysis of injuries found that head, face, and neck injuries accounted for ∼3.6% of all rowing-related injuries.^34^ Due to the limited amount of data available within the sport, the knowledge surrounding the lasting impact of SRC in rowing is also unknown.

### Cross Country and Track and Field

While not typically considered a sport in which SRC routinely occur, it has been reported that high schools cross country runners had an 18.7% chance of sustaining a SRC while, more generally, track and field had a 22.8% chance.^35^ Most SRC occur due to contact with the ground, from falls and loss-of-footing generally and especially but not exclusively during vaulting or jumping events^36^. Both female track and field and cross country had more recorded SRC than their male counterparts. While both sports are relatively low-risk, concussive events and potential sudden deaths have been reported. ^37^ Track and field, including cross-country, would benefit from a formal effort to record more carefully the mechanism(s) of injury and in each type of sport and the risks of head injury.

### Diving and Swimming

While rare, SRC occur within these aquatic sports, notably springboard and platform diving and repetitively hitting the water at high speeds. In a review of high school aquatic sports from 2013-2018, it was found that female swimmers sustained an incidence rate of 0.66 per 1,000 AE, which is two-fold the risk of male swimmers and divers 0.37 per 1,000 AE. ^40^ From the same study, it was found that females reported 5.6% of SRC sustained to be recurrent from those small number of injuries.^40^ These findings additionally found that female swimmers and divers were nearly as likely as male soccer players to have a recurrent SRC (5.6% vs. 5.7%).^40^

### Equestrian

The equestrian sports continue to grow in the United States and internationally, with 41 female participants representing the majority of 10,000 athletes competing in U.S. collegiate equestrian sports yearly.^25^ Not surprisingly, head injuries are pretty common; horse riders can expect a severe accident once every 350 hours of participation, which is twenty times more frequent than motorcycling.^25^ Indeed, patterns of injury in equestrian sports found almost 50% of injuries included neurological insult.^42^ Side-by-side comparisons of horseback riding and hockey observed that horseback riding-related injuries were more likely to require hospitalization.^43^ Furthermore, it has been found by several studies that equestrian activities likely have higher mortality rates than football, rugby, motor racing, and skiing. ^44^

### Field Hockey

Field Hockey is a high contact sport and is considered an extremely high paced and intense sport where the risk for sustaining a concussive injury can be high.^45^ SRC account for the majority of all injuries within this sport,^47^ but there is extraordinarily little detailed injury statistics available.^29^ Recently, Kerr and colleagues tried to bridge this gap by investigating girls’ high school and collegiate field hockey players’ and found SRC risk was similar to that found in Division I American football players.^2^ A more recent study, surveyed the injury incidence in international field hockey, considering both sexes, and reported that the most common injury was to the face and head.^48^ Although mechanism was not reported within this specific study, previous research has suggested the most common injury mechanisms were associated with making contact with another player, or by being hit with a ball or stick.^46^,

### Lacrosse

Boys lacrosse is a full-contact sport allowing the body stick contact (e.g. “checking”), mandates that hard shell helmets with full face masks be worn at all times.^50^ Whereas, in girls’ lacrosse, body checking is not allowed, and certain rules are supposed to prevent stick checking to the head. However, flexible headgear with/without integrated eye protection is not always required.^50^ Since the 1950’s, women have sustained higher percentages of head and facial injuries occurred form stick or ball contact in contrast to athlete-athlete contact.^51^ In a recent study, it was found that boys lacrosse players sustained 0.47 SRC per 1,000 AE while girls lacrosse players sustained 0.39 SRC per 1,000 AE.^50^ Another analysis of injuries uncured during competitions found similar findings female athlete sustained 0.83 SRC per 1,000 AE compared to males who only sustained 0.56 SRC per 1,000 AE.^52^ The limited literature and available data suggest that improved reporting and documentation of head injuries in lacrosse is warranted.

### Luge

While luge competitions have taken place since the mid-1800’s, it was not until the late 1900’s that coaches became concerned about athletes almost universally complaining about headaches and other symptoms related to head injuries. Some symptoms were reported as lasting only minutes, others, for days; as the number of runs increased, so did the likelihood and severity of headaches.^53^ Reports of, so-called, “sled head,” described as headaches, fogginess, and occasionally disequilibrium resulting mostly from bumpy or multiple tracks runs became common.^54^ Headaches presented bilaterally and were described as “throbbing” or “constant”. Headaches reportedly progressed in severity throughout a day of training, limiting the number of runs an athlete could perform,^53^ the jolting motion of the toboggan at high speeds, for long durations, can be the cause of these symptoms and be the mechanism for which SRC occur. As an Olympic sport, it might be expected that more data might be available, however, a paucity of information exists on modern record keeping.

### Softball

Long popular, girls’ and women’s’ softball enjoys roughly 10-to-12 million athletes participating annually in the US. Many elements of softball play involve high-energy events that can cause head and facial injuries, including pitching, batting, or running into other players or objects.^55^ In one recent study in looking at bothyoung and adult female athletes, head injuries and SRC accounted for 45.0% and 17.7% of all injuries, respectfully. This study also estimated that emergency departments in the United States see 121,802 softball-related head and facial injuries per year.^55^ In two large cohort studies, high school and collegiate athletes’ SRC injury risk was reported as 0.07 per 1,000 AE (56, 57) additionally, both studies reported significantly greater proportion of total concussive injuries in softball plyers than in baseball players (5.5% and 2.9% respectively).^2^

### Snowboarding and Skiing

Head injuries constitute 16%–27% of injuries among children and adolescents engaged in snow sports, such as alpine skiing and snowboarding, with over half of these injuries may be classified as traumatic brain injuries (TBIs), including SRC, cerebral contusions, and intracranial hemorrhages.^58^ In a 2002 review it was reported that youths had the highest risk for a head injury at 10-13 years of age (1.69 per 1,000 participants).^60^ Following these results and increasing participation within both sports, more in-depth investigation has been done to try to understand these risk trends. Investigation of emergency department visits in youths for snow sport-related TBIs from the year 1996-to-2010 indicated that out of >700,000 injuries, 78,538 were classified as head injuries. ^58^ Even though both sports are vaguely investigated, the reported incidence rate of 2.11 SRC per 1,000 AE was higher than most studies involving soccer and even football. ^58^

### Surfing

Surfing has a long history as a recreational and competitive sport - one designated as an Olympic sport in 2020. Though an aquatic sport, 91% of surfers admitted to sustaining a sport-related injury at some point during their career. ^62^ The mechanism of injury is poorly documented, but most surfers reported having accidental impact with the surfboard during a hard fall, colliding with another surfer, or contacting rocks, coral, or beach.^61^ It has been suggested that the annual estimate of surfing-related head injuries increased from 1,189.71 in 2001 to 1,790.66 in 2016. ^62^ Due to surfing being a highly recreational sport, it is important those involved understand the risk, symptoms and reporting policy when getting injured in the water. Without proper SRC protocols and education for surfers, lifeguards, as well as those in charge of surfing competitions, thereby improving reporting, it will be difficult to ever understand the true risk of injury.

### Tennis

SRC in tennis are extremely rare, although, players have experienced being hit in the head with a ball during play, taken a fall, or have been hit in the head with a tennis racket.^64^ Pueringer et al, searched *The National Electronic Injury Surveillance System* on reported past adult tennis-related head and facial injuries from the previous 10 years and found that around 11% of those injuries were classified as a SRC.^36^

### Volleyball

In spite of volleyball being a growing and intense sport, USA Volleyball is one of the few organizations that does not have an official SRC protocol or training requirement.^66^ Reeser et al. investigated the incidence of volleyball-related head injuries between among both high school and NCAA athletes, finding that SRC represented one of the top five diagnoses and the third most frequent in the high schoolers.^67^ Additionally, they found that just over half the competition-related SRC in both college (56.0%) and high school (60.9%) occurred while digging (diving for the ball before it hits the ground) and making a defensive play.^67^ Similar findings analyzing collegiate women’s and men’s volleyball players discovered that most SRC were due to ball contacting the head (men, 83.3%; women, 53.6%),^68^ SRC were the second most common injury among both groups behind spraining a wrist or ankle. Even though volleyball is not as high-risk for SRC as in soccer, for instance, their presence in a limited-contact sport highlights the need for continued targeted SRC research and prevention efforts.^67^

### Water Polo

Water polo is the only sport where at all levels there is no standardized set of procedures for reporting SRC and is the only NCAA-sanctioned sports in which an injury database is not available.^69^ Therefore, in 2016, in a first-of-its-kind study, Blumenfeld et al. surveyed water polo collegiate male athletes who reported sustaining an average of 2.27 “serious blows to the head” imposed by both the ball and opposing athletes per game over the span of their career.^70^ One of the few investigations conducted found the within the youth population mean number of what they considered “blows to the head” during practice was 1.85±0.08 and 2.27±0.07 during a game. ^71^ Water polo is assumably at higher-risk for sub-concussive contact or, given its association with other lower-risk aquatic sports, under-reporting. As the most high-contact or low-contact aquatic sports, water polo may present as a particular example of head-injury risk in water sports, generally.

## A META-ANALYTIC ASSESSMENT OF STUDY FACTORS ON REPORTED INCIDENCE RATES

The rates of SRC vary widely per sport, often due to factors or how the sport is played but also to factors outside of the game itself. In many ways, however, the rates of SRC concerning these and other sporting activities appearing in the scientific literature are influenced not by events happening on the field, court, during practice or competition, but rather are related to how the studies were performed and the characteristics of those published research articles. Meta-analytic approaches are particularly valuable on their own or as a complement to a narrative review by statistically highlighting variables which influence or modify factors on reported study results. Such analyses not only offer the means to estimate population-level effects but also give insights into the sociological factors of how the science was conducted, analyzed, and reported. Quantifying such factors can assist future studies by ensuring best practice, optimizing sample characteristics, and minimizing the effects of unintended biases.

In the following section, a modest, but likely among the first, meta-analytic investigation is presented to explore the study moderator variables which contribute to the variance of incidence reporting rate effect sizes in the literature. Due to a lack of data on recreational or travel sports, the whole population of interest for our quantitative analysis was youth or high school studies that looked at both males and females in sex-comparable school-sanctioned sports (Baseball, Softball, Cross Country, Basketball, Cheerleading, Gymnastics, Cricket, Crew, Field Hockey, Lacrosse, Swimming/Diving, Tennis, Track and Field, and Volleyball). This analysis aims to identify the study factors contributing to incident differences in peer-reviewed research publications. Different methods by which data is collected, lack of reporting standards, and difficulty in identifying causal relationships between SRC and incidence risk is a promising area for meta-analytic examination.

## METHODS

### Studies Included

From 1989-2021, a total of 108 studies were identified through PubMed and Google Scholar; these databases were searched using keywords such as ‘Sports-related Concussion,’ ‘Athletic Exposures,’ ‘Neuroimaging,’ and ‘Return-to-Play.’ Due to the lack of cohesive experimental designs between neuroimaging studies, there was an inability to model the results to accurately make conclusions statistically. Thus, the incidence rate became the primary focus of this systematic review. 57 studies met inclusion criteria for the qualitative analysis, and of those 57 studies, 15 had quantifiable incidence rates in units of athletic exposure by 1000 hours. (**Figure 1**)

**Figure 1:**
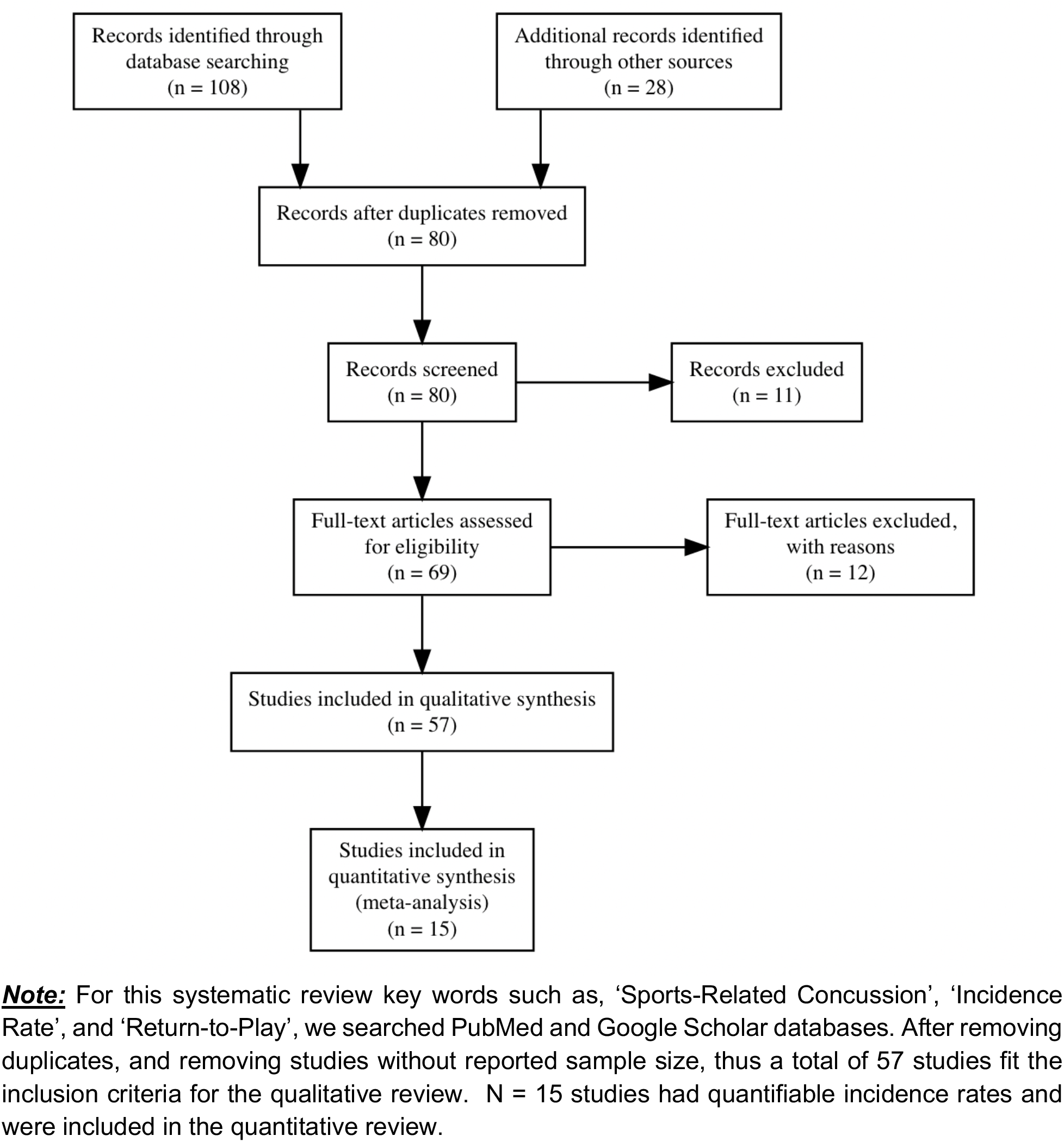
PRISMA Meta-Analysis Exclusion and Inclusion Flow Chart

We identified N=15 of those studies to have quantifiable athlete exposure (AE). This was defined as a single student-athlete participating in a single sanctioned practice or competition in which he or she was exposed to the possibility of athletic injury, regardless of the time associated with that participation. After removing previously well-studied sports, (e.g., Football, Soccer, Ice Hockey, and Wrestling), a total of 84 data points, including reported incidence rates from the 15 peer-reviewed literature, were computed (**Table 2**). All rates were re-scaled to AE units as one athlete participating in one practice or competition during which the athlete was exposed to the possibility of athletic injury, against a baseline of 1,000 AEs.

**Table 2.** Studies of Sports-Related Concussions (1988-2022) Please see attached spreadsheet file

**Table 3.** Selected Studies for Systematic Review of Concussion Research from 1988-2019

| Citation | Sport | Sex | Number of Authors | Year of Publication | Duration of Study (yrs) | Years Studied | Studied Started Before 2010 | Study Sample Size | Mean Incidence Risk (SD) |
| --- | --- | --- | --- | --- | --- | --- | --- | --- | --- |
| Agel et al<sup>72</sup> | Basketball | F | 7 | 2007 | 6 | 1998-2004 | Yes | 10,221 | 0.34(0.00) |
| Allen et al<sup>17</sup> | Basketball | F/M | 7 | 2019 | 3 | 2011-2014 | No | 167 | 0.35(0.13) |
| Baugh et al<sup>49</sup> | Volleyball | F/M | 6 | 2018 | 2 | 2014-2015 | No | 18,844 | 0.37(0.03) |
| Gessle et al<sup>2</sup> | Multisport | F/M | 5 | 2007 | 2 | 2005-2006 | Yes | 396 | 0.21(0.15) |
| Hill et al<sup>19</sup> | Cricket | F/M | 5 | 2021 | 4 | 2015-2019 | No | 485 | 0.40(0.00) |
| Kerr et al<sup>16</sup> | Multisport | F/M | 7 | 2017 | 2 | 2015-2016 | No | 73 | 0.56(0.71) |
| Kerr et al<sup>81</sup> | Multisport | F/M | 6 | 2020 | 6 | 2013-2018 | No | 9,542 | 0.52(0.40) |
| Lincoln et al<sup>30</sup> | Field Hockey | F/M | 5 | 2007 | 4 | 2000-2003 | Yes | 8,638 | 0.19(0.15) |
| Lincoln et al<sup>59</sup> | Multisport | F/M | 6 | 2011 | 11 | 1997-2008 | Yes | 2,651 | 0.24(0.05) |
| Lynall et al<sup>47</sup> | Field Hockey | F | 11 | 2018 | 6 | 2008-2014 | Yes | 1,979 | 0.36(0.00) |
| Mara et al<sup>38</sup> | Multisport | F/M | 4 | 2012 | 3 | 2008-2010 | Yes | 4,635 | 0.54(0.34) |
| Marshall et al<sup>74</sup> | Softball | F | 5 | 2007 | 18 | 1988-2004 | Yes | 5,336 | 0.16(0.00) |
| O'Connor et al<sup>36</sup> | Multisport | F/M | 6 | 2017 | 4 | 2011-2014 | No | 2,004 | 0.29(0.26) |
| Reeser et al<sup>8</sup> | Volleyball | F | 4 | 2015 | 4 | 2005-2009 | Yes | 1,979 | 0.13(0.00) |
| Rosenthal et al<sup>39</sup> | Multisport | F/M | 5 | 2014 | 7 | 2005-2012 | Yes | 4,024 | 0.35(0.28) |
| **TOTALS** |  |  | **89 authors** | **2007-2021** | **82 cum. years** | **1988-2019** | **9/15 studies begun prior to 2010** | **70,974** | **0.34(0.36)** |

**Note.** Studies here met inclusion criteria for meta-analytic assessment. Impact factors per study are sport, sex, number of authors, year published, duration of study, years studied, whether the study started before the 2010 CDC Rule Change and the estimate population sample size. SD < 0.001 are reported here as zero.

**Table 4.** Study Descriptive Statistics before Removing High-Exposure Sports

| Predictor Variable | n | Mean | SD | Median | Skew | Kurtosis | SEM |
| --- | --- | --- | --- | --- | --- | --- | --- |
| Incidence Rate | 115 | 0.33 | 0.36 | 0.22 | 2.88 | -13.2 | 0.03 |
| Number of Authors | 115 | 5.65 | 1.05 | 6.0 | 0.77 | 4.23 | 0.10 |
| Year Published | 115 | 2014 | 3.88 | 2017 | -0.63 | -0.82 | 0.36 |
| Duration of Study | 115 | 4.85 | 2.93 | 4.0 | 1.55 | 2.77 | 0.27 |
| Studied Started before 2010 | 115 | 1.53 | 0.50 | 2.0 | -0.12 | -2.00 | 0.05 |
| Sample Reported Injuries (N) | 115 | 3738.32 | 2858.78 | 262.033 | 1.29 | 0.21 | 266.58 |
| Concussions per Sport (n) | 115 | 258.58 | 1048.62 | 7.0 | 7.96 | 69.63 | 97.78 |
| Proportion of Females | 115 | 56.00 | 34.73 | 1.4 | -0.41 | -1.01 | 3.24 |

**Note:** Descriptive Statistics were computed for all included variables. As seen above, 56% percent of recorded concussions pertains to female athletes. An overall average incidence rate for both females and males was additionally computed and discovered to have a large standard deviation implying data has a large observed variance ( $m = 0.33$ , $s = 0.36$ )

For the purposes of this analysis, youth and college-level ages were exclusively studied, emphasizing what influences reporting rates within these two populations. Many of the studies used a retrospective longitudinal approach by quantifying exposure risk in various school-sanctioned male and female sports concurrently. Female and male athletes were not separated within the data and sex was used as an independent variable. The targeted age was 5-18, therefore using high school injury surveillance to distinguish an understanding of the youth population. Sports with less than two studies were also excluded. Additionally, a discrepancy was found within reported sample sizes, as some investigators classified the number of injuries as a sample size, but the number of participants was not registered. Therefore, due to the purposes of this analysis and unless reported otherwise, those with reported injuries or reported sample size were considered the same.

**Table 5.** Study Descriptive Statistics after Removing High-Exposure Sports

| Predictor Variable | N | mean | SD | Skew | Kurtosis | SEM |
| --- | --- | --- | --- | --- | --- | --- |
| Citation | 85 | 9.26 | 3.60 | -0.25 | -0.98 | 0.39 |
| Incidence Rate | 85 | 0.23 | 0.23 | 1.68 | 3.25 | 0.02 |
| Number of Authors | 85 | 5.66 | 1.10 | 0.96 | 4.85 | 0.12 |
| Year Published | 85 | 2014.79 | 3.84 | -0.69 | -0.74 | 0.42 |
| Duration of Study | 85 | 4.81 | 2.93 | 1.79 | 3.95 | 0.32 |
| Studied before 2010 | 85 | 1.55 | 0.50 | -0.21 | -1.98 | 0.05 |
| Population of Injuries (N) | 85 | 3633.79 | 2936.18 | 1.29 | 0.17 | 318.47 |
| Concussion per Sport (n) | 85 | 208.00 | 1123.25 | 8.38 | 71.36 | 121.83 |
| Proportion of Females Concussed | 85 | 65.06 | 30.63 | -0.61 | -0.41 | 3.32 |

**Note:** Once removing Football, Soccer, Ice Hockey and Wrestling, descriptive Statistics were computed for all included variables. As seen above, an increase of 65% percent of recorded concussions pertains to female athletes. Yet, an overall lower average incidence rate for both females and males implied data had a smaller observed variance ( $m = 0.23$ , $s = 0.23$ ).

**Table 6.**
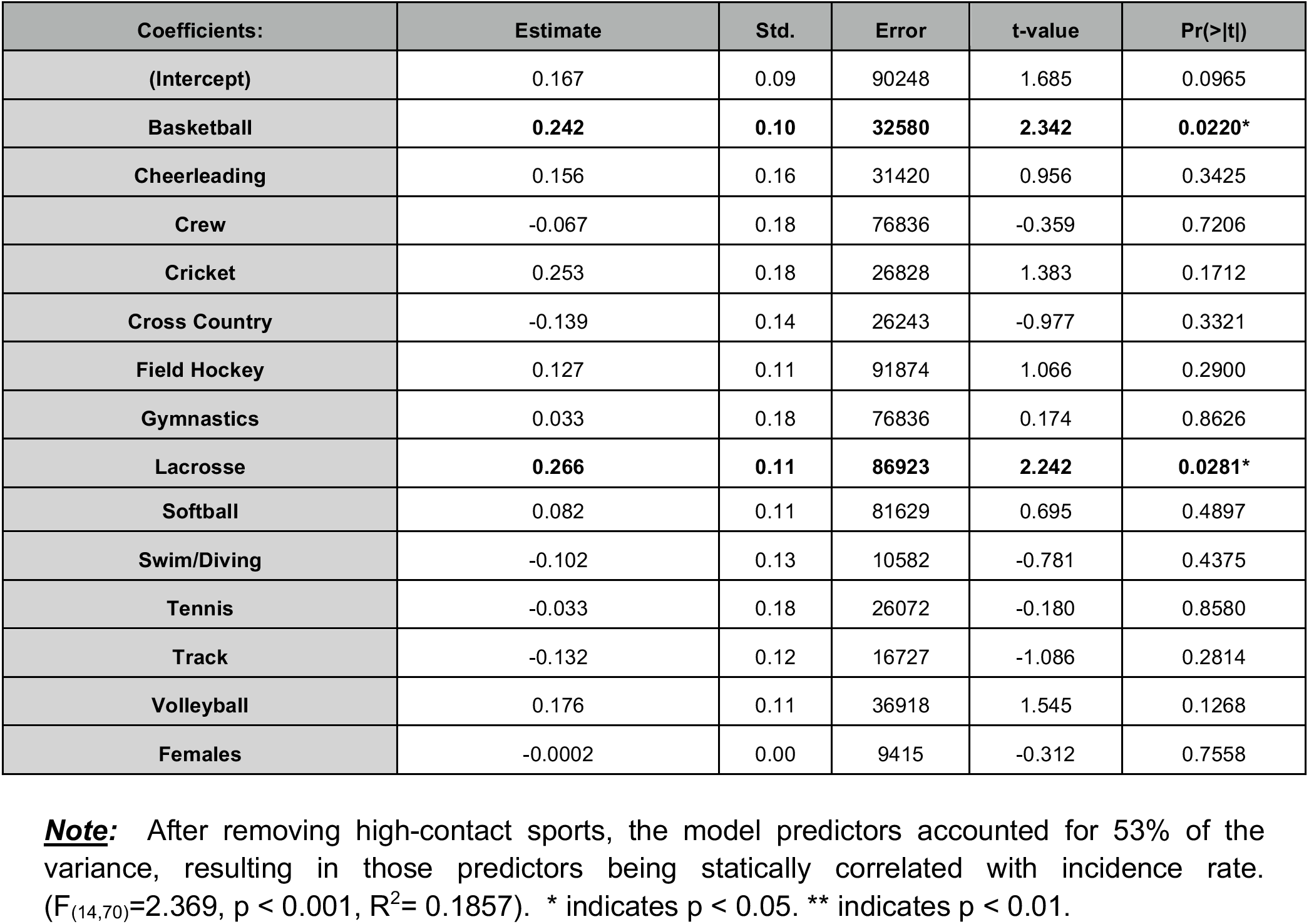
Regression Model Parameter Estimation

### Specific Study Modulating Factors

As seen in **Table 1**, study variables encoded from each of the included articles were identified as:

#### Sport

For each sport, for each study, the presence or absence of that sport was dummy coded such that, in included, it was coded as unity, and, if not, it was coded as zero. In multi-sport incidence reports, multiple studies might be present, with the presence of each being denoted.

#### Study Sample Size

The sample size of a study is often used as a proxy for overall study quality. In effect, studies including a greater number of subjects tend to be better designed, controlled, and are more robust.

#### Proportion of Females

The proportion of females contributing to the total number of SRC reported by each study was determined. Thus, where a study reported the results of sports which included all females, the value would be 100%. Conversely, “male sports” would be coded with a value of 0%. Studies which included both males and females in their reported incident rate results would have values ranging from 0-100%.

#### Year of Study Publication

The year in which the study appeared in the peer-reviewed literature was used to help assess secular trends in SRC incidence rates over time.

#### Duration of Study

The period in years over which the study was conducted. Longer study periods may be an index of research support and infrastructure, each of which could influence the reported incidence rates.

#### Studied before 2010

If the legislation influenced SRC rates in the wake of the Lystedt Law (September 2010), after which states in the US, the CDC, and other organizations began to more strictly enforce sports SRC regulations. A causal relationship between higher SRC incidence rates and heightened SRC awareness has been inferred by previous researchers but, has yet to be established.

#### Number of Contributors/Authors

The number of authors is used in meta-analytic assessment as a further indicator of study quality since, presumably, the more authors there are, the more rigorous the work might have been performed due to the size of the research team and the number of individuals contributing to the final published report.

### Power Analysis

Heterogeneity is regularly considered when defining the variability between studies. When defining heterogeneity, Higgins and Thompson^65^ identified *I*^2^ values to determine the difficulty in drawing overall conclusions from retrieved studies. When *I*^2^ is near or close to zero, it is assumed the observed variability is mostly due to sampling error. Thus, when *I*^2^ is closer to 100, most of the observed variability reflects differences in population effect sizes. Due to high differences in the population sampling sizes, a moderate *I*^2^ of 50% was used. This index is a useful tool for the degree of heterogeneity across studies when the null hypothesis of having less variability than expected from chance is rejected at an α < 0.10. A mixed-effects power analysis model to computed to summarize the effect size estimate for the two established groups, those whose incidence rates are above the mean and those who reported below the mean. For the analysis to have 80% power to detect an effect of 20% (d = 0.20), with a heterogeneity estimate of *I*^2^ = 0.05, 70 participants were required in each of the two groups. The final sample satisfied those requirements.

### Multilevel Regression

General linear models (GLM) were used in two separate analyses to examine the influence of specific sport and sex on the incidence rate of SRC, and to examine the influence of study characteristics on reported incidence rates. Multilevel linear regression was chosen for a few two specific reasons, the main goal was to obtain full item information and to further estimate the relationship between the modulating factors of the research can successfully predict the linear effect of the AE incidence mean value. Additionally, despite missing data, imputations were not considered, thus multilevel linear regression this model assumes the predictor variable (Incidence Rate) will generate a regression equation, the equation is then used to predict these missing values. To further estimate the most influential predictors within the nested meta-analytic review, backward stepwise linear regression was then used to identify any possible predictors of the incidence rate. At each step, variables were removed based on the p-values. Therefore, the variables with the lowest correlation were removed. Statistical significance was set at p < 0.05. All analyses were conducted using the R Statistical Programming Environment.^68^

### Regression within Sport and Incidence Rate

This analysis explicitly examined the influence of specific sports on the incidence rate in relation to the proportion of females concussed. Two models were. To fully understand this influence within the different sports, and to avoid the overshadowing effect of sports with high exposure rates, sports including Football, Soccer, Ice Hockey, and Wrestling were explicitly removed from the analysis. A basic linear regression without backward elimination was used (Equation 1). Finally, an independent Student’s t-test was conducted on the overall proportion of SRC reported between females and males in their corresponding sports.

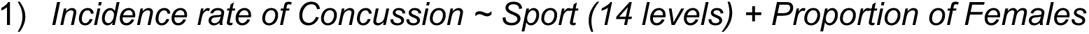

### Characteristics of Research Studies on Reported Incidence Rates

To investigate the influence that specific characteristics from these studies have on the reported SRC incidence risk a second GLM analysis was performed. The characteristics included, the duration of the study, the proportion of females concussed within that sport, the number of authors and contributors, the year the article was published, total reported injuries, and lastly, whether the article was published before or after the 2010 legislation (Equation 2). Two models were constructed to understand the influence that male-dominant, high-exposure sports had an influence on the incidence rate, specifically within the proportion of females concussed. Therefore, one model contained all sports, whereas model two removed Football, Soccer, Ice hockey and Wrestling. A variable coding pre-/post-2010 was included because previous findings have suggested that passage of the Lystedt Law has resulted in increased awareness and legal requirements leading to increased reporting of SRC, and thus greater reported SRC risk.

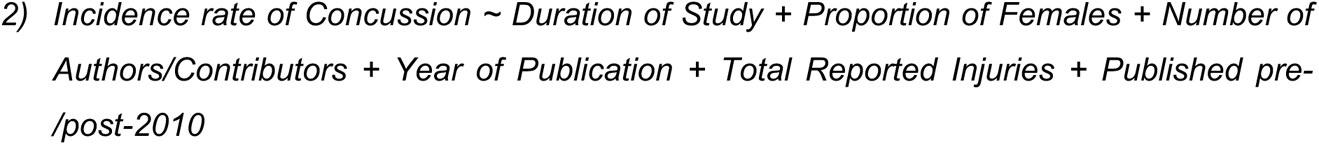

## RESULTS

### Regression within Sport on Incidence Reporting

The results of the regression indicated that the model was able to significantly predict incident rates for SRC (F_(14,70)_=2.369, p < 0.001, R^2^= 0.1857). The proportion of females that were concussed was not a significant predictor of the incidence rate. Surprisingly, Basketball (b=0.24, t=2.342, p<0.02) and Lacrosse (b=0.27, t=2.242, p<0.03) both were significant indicators. While lacrosse can be known as a high-contact sport, basketball isn’t, and therefore has less research exposure than lacrosse. These results suggest that in studies that include basketball with either female or male athletes, may be a significant predictor of a higher reported incidence rate.

A Student’s t-test was conducted to investigate the difference in proportional differences in sustaining a SRC between females and males based on their sport. Results of the independent sample t-test indicated that there was a significant difference between the proportion of sustained SRC between females and males (t_111_=4.702, p<0.001). The overall proportion of females to males concussed was 69.2% female vs 40.9% male, with an overall large effect, d=0.879. These results may indicate that studies are collecting more female SRC per sport, yet this larger percentage SRC was not correlated with a higher reported incidence risk. Thus, studying both females and males in congruent sports, and within the same study, may influence the reported female incidence rate.

**Table 7.** Multiple Linear Regression modulating impact factors within concussion research

| Coefficients: | Estimate | Std. Error | t-value | Pr(>\|t\|) |
| --- | --- | --- | --- | --- |
| (Intercept) | -0.06 | 0.13 | -0.44 | 0.66 |
| Number of Authors | **0.06** | **0.02** | **-2.95** | **0.001\*\*** |
| Duration of Study | **-0.02** | **0.008** | **-2.08** | **0.04\*** |

**Note:** Backward elimination method identified the two significant modulating impact factors as number of authors and duration of study ( $F_{2,82}=5.93$ , $p<0.001$ , $R^2=0.1051$ ). The number of authors positively affects to incidence rate, while the longer the duration of the study the higher association a lower incidence risk. \* indicates $p < 0.05$ . \*\* indicates $p < 0.01$ .

### Multiple Linear Regression on Study Characteristics on Reported Incidence Rates

The results of the multiple regression indicated that there were significant nested effects between the number of authors in addition to the duration of the study (F_2,82_=5.93, p<0.001, R^2^=0.1051). Backward elimination further investigated these individual predictors, and a stepwise approach accounted for all variables that were deemed, insignificant predictors. The number of authors (b=0.06, t=2.95, p<0.001) was the only positive indicator of incidence rate, meaning, for every author added there was a 0.06 increase in that study’s overall incidence rate. Duration of study (b=-0.02 t=-2.07, p<0.05) was a negative indicator of incidence risk, indicating that, for every year added to the study, there was a decrease in incidence rate by 0.02.

Before removing male-dominant high-exposure sports, our model indicated that the proportion of females did significantly affect the incidence rate (b= −0.003, t= −4.22, p< 0.001). Once removing these sports, the model demonstrated that the proportion of females concussed did not statically affect the incidence rate (p =0.27). This indicated that the possibility that including female sports in analyses which include these high-exposure sports, may influence the incidence reporting rate.

## DISCUSSION

Identifying those athletes and the sports in which they participate which are at greater risk for sport-related concussion (SRC) and/or prolonged recovery represents an important element for informing prevention efforts and management approaches. Importantly, coaches, trainers, and clinicians who are aware of factors that influence SRC risk and recovery are more informed and better prepared to discuss diagnosis, prognosis, and expectations for recovery with the injured athlete.

After controlling for the effects of well-studied sports that are already the focus of SRC prevention efforts, meta-analytic results illustrated that lower profile sports, such as Basketball and Lacrosse, have significant, positive relationships with higher incidence rates. The relationship between the number of authors and results has yet to be fully investigated, but as the results from this systematic review demonstrate, studies that contain a large number of authors could potentially have an influence on overall results. Large research consortia seek to pool data from across multiple sites and, perhaps, multiple sports, whereas smaller study teams may only be reporting the results from a single site and only a single sport. More authors also may have a mitigating effect on reported incidence rates due to more input on the final results and the resulting research article. Thus, large-scale efforts involving multiple study contributors may tend to under-report SRC incidence.

Conversely, smaller studies reporting significant effects relative to low sample sizes may expedite their results to publication. In such cases, smaller studies may tend to overestimate reported SRC incidences. This phenomenon is often referred to as *publication bias*, affecting studies’ reliability in many segments of the literature. It is likely to also influence reported SRC rates from sports-related activities.

Additionally, longer-duration studies, spanning more than four years at a time, were correlated with a lower incidence rate reporting. One limitation to collecting longitudinal retrospective data outside a formal study is that researchers cannot be sure the same participants are being evaluated the same during each separate hospital visit. This can lead to an underestimation of variability and an increased likelihood of Type II statistical error. For example, several studies examined here reported incidence rates reliant on the *National Athletic Treatment, Injury and Outcomes Network (NATION)* – a database established in 2011 intended to devise a comprehensive injury surveillance system for high school students. Despite being a valuable tool for investigators, the data contained in NATION are retrospectively collected, which may have resulted in certain ascertainment biases and under-reporting of actual SRC events at the time of injury.

In youth, high school, as well as collegiate athletics, the participation of female athletes has increased dramatically since the implementation of Title IX as part of the noteworthy Equality in Education Act of 1972.^2^Due to this increase in participation, the annual incidence of sports-related SRC can be expected to increase, with female athletes at higher injury risk than concussed male athletes. As this review found, female athletes’ presence modulates incidence risk, but whether females are experiencing and reporting higher SRC risk due to physiological sex differences or externally related factors has yet to be established.

As mentioned, SRC injury-risk and symptom behaviors has been difficult to consider meta-analytically as most of the literature did not report these variables in a sufficient manner suitable for external or generalizable analyses. This might not be surprising given a recent bibliometric analysis which identified the top one-hundred cited sports concussion articles noting that injury prevalence was the dominant reported metric whereas treatment efficacy was only considered in lesser cited articles^79^. Despite SRC meta-analytic reviews, such as this one, having suggestively low in power, these reviews can be particularly helpful in assessing the structure in which current clinical data is being collected and processed. Furthermore, a universal data repository on SRC injury risk, recovery time, and other factors would be an effective way to cohesively work with other researchers in the field to understand the modulating factors surrounding SRC effectively.

The studies included in this meta-analysis suggested that females were not experiencing higher incidence rates of SRC compared to males. Rather those included female athletes that were analyzed within a large group setting with high-exposure sports (e.g., Football, Ice Hockey, etc.), resulting in a higher proportion of concussed females within a single study not significantly correlated with having an impact on the study’s population incidence rate. These results suggest that the actual incidence rate of females may be modulated by external factors (e.g., high-contact sports, gender influences, research protocol, socioeconomic and socio-environmental differences), in contrast to a higher risk of injury as the current literature suggests.

## CONCLUSION

Sports-related concussion research has gained considerable attention, and it is essential to understand the risk of SRC beyond high profile and high contact sports and the effect of sex. The result from this quantitative meta-analysis suggests that, in a subset of adolescent sports, there is no significant predictive effect of sex, despite a significantly higher proportion of SRC diagnoses in female athletes. This meta-analysis also explored the frequency and means of SRC beyond high-contact sports and found that features of SRC studies can have significant effects on reported rates of trauma. Factors such as these should be considered when critically evaluating research on the care and risk of SRC for both males as well as for female athletes. This study suggests that studying both females and males in congruent sports and within the same survey, maybe resulting in influencing reported rates for female athletes. Further investigation is likely needed into socio-environmental and socioeconomic issues affecting female athletes’ medical care. Finally, moving forward, comprehensive, national tracking databases, where SRC are recorded across sporting activities, institution/organizations, for men, women, and so forth, would be advantageous for subsequent analyses of SRC due to athletic exposures.

## Data Availability

All data produced in present work are contained in the manuscript

## ACKNOWLEDGEMENTS

R.E. and J.D.V.H. conceived of the study, R.E. conducted the literature review, gathered data on study factors, conducted statistical analyses, and drafted the manuscript. J.D.V.H. contributed to the editing of the final draft manuscript. The authors wish to thank Dr. Benjamin Newman for critical comments on early drafts of this manuscript.

## Footnotes

1 Washington State Legislature, RCW 28A.600.190, *Youth sports—Concussion and head injury guidelines—Injured athlete restrictions—Short title*. https://apps.leg.wa.gov/rcw/default.aspx?cite=28A.600.190

2 See 20 U.S.C. Ð 1681 – 1688, TITLE 20 – Education, CHAPTER 38 - DISCRIMINATION BASED ON SEX OR BLINDNESS, US Department of Justice, https://www.justice.gov/crt/title-ix-education-amendments-1972.

