## Supplementary material for "Modulating Factors Affecting Sports-Related Concussion Exposures: A Systematic Review and Analysis": Table 2

| Sport | Title | Authors & Coauthors | Research Area of Focus | Journal Published | Year Published | # Authors | Sample Size Reported | Years studied | Type of Study | Country Performed | Goal of Research |
|---|---|---|---|---|---|---|---|---|---|---|---|
| Volleyball | Injury Epidemiology and Time Lost From Participation in Women's NCAA Division I Indoor Versus Beach Volleyball Players | Tristan Juhan, Ioanna K Bolia, Hyunwoo P Kang, Andrew Homere, Russ Romano, James E [illegible] | Concussion Prevalence | Orthopedic Journal of Sports Med | 2021 | 8 | 161 (53 beach 108 outdoor) | 2015-2016 | Cohort study | USA | To compare the epidemiology of injuries and time lost from participation between female NCAA Division I athletes who participate in [illegible] |
| Basketball | Concussion incidence and Return-to-play time in National Basketball Association Players | Ajay S Padaki, Brian J Cole, Christopher S Ahmad | Concussion Prevalence | American Journal of Sports Medicine | 2016 | 3 | 315 | 2006-2010; 2011-2014 | Descriptive Epidemiological Study | USA | The incidence and return-to-play statistics were generated by synthesizing information from publicly available records |
| Equestrian | Horse-related incidents and factors for predicting injuries to the head | Lauren Meredith, Robert Ekman, Robert Thomson | Concussion Prevalence | BMJ Open Sport & Exercise Science | 2018 | 3 | 7815 | 2001-2016 | Retrospective Review | Sweden | Investigate the association between incidence type and head injury in equestrian-related sports. |
| Equestrian | Concussion History and Knowledge Base in Competitive Equestrian Athletes | Heather N Kuhl, David Ritchie, Angela C Taveira-Dick, Katie A Hoefling, Stephen A Russo | Concussion Prevalence | Sage Journals | 2013 | 6 | 94 | 2010 | Cross-sectional level 2 | USA | Concussions per rider |
| Equestrian | Adult sports-related traumatic brain injury in United States trauma centers | Ethan A Winkler , John K Yue, John F Burke , Andrew K Chan, Sanjay S Dhall, Mitchel S Berger, Geoffrey T [illegible] | Concussion Prevalence via Hospital/Urgent Care Records | Neurosurg Focus | 2016 | 8 | 4788 | 2003-2012 | Retrospective | USA | Utilizing the National Sample Program of the National Trauma Data Bank (NTDB), the authors retrospectively analyzed sports-related TBI data from adults (age ≥ 18 years |
| Football | Concussions From Youth Football: Results From NEISS Hospitals Over an 11-Year Time Frame, 2002-2012 | Nathan A. Jacobson, David Buzas, Lawrence G. Morawa | Concussion Prevalence via Hospital/Urgent Care Records | Orthopedic J Sports Med | 2013 | 3 | 2028 | 2002-2012 | Descriptive Epidemiological Study | USA | Over an 11-year span from January 2002 to December 2012, the authors reviewed the concussions sustained by ath- letes aged 5 to 13 years while playing football, as evaluated in emergency departments (EDs) in the United [illegible] |
| Gymnasstics | Health outcomes among former female collegiate gymnasts: the influence of sport specialization, concussion, and disordered [illegible] | Emily Sweeney, David R Howell, Corrine N Seehusen, David Tilley, Ellen Casey | Concussion Prevalence | The Physician and Sports Medicine | 2020 | 5 | 437 | Not Reported | Survey | USA | Participants were put into two groups, (<14 years) or late (> or equal to 14 years) specialization, in addition to whether they [illegible] |
| Horse Riding | Protective and risk factors in amateur equestrians and description of injury patterns: retrospective data analysis and case-control survey | Rebecca M Hasler, Lena Gyssler, Lorin Benneker, Luca Martinolli, Andreas Schötzau, Heinz Zimmermann,and Aristomenis K Exadaktylos | Concussion Prevalence via Hospital/Urgent Care Records | Journal of Trauma Management & Outcomes | 2011 | 7 | 365 | 2000-2006 | Retrospective / case control survey | Switzerland | To analyse injury patterns, protective factors, and risk factors related to horse riding, and to define groups of safer riders and those at greater risk |
| Luge/Skeleton | Concussions in Sledding Sports and the Unrecognized "Sled Head": A Systematic Review | Melissa D McCradden, Michael D Cusimano | Concussion Prevalence Systematic Review | Frontiers in Neurology | 2018 | 2 | 40 | 1946-2017 | Systematic Review | Canada | To summarize our knowledge of the prevalence of concussion and related symptoms in sledding sports; |
| Snowboarding/skiing | Head Injuries in Skiers and Snowboarders in British Columbia | S Hentschel, W Hader, M Boyd | Concussion Prevalence via Hospital/Urgent Care Records | The Canadian Journal of Neurological Sciences | 2000 | 3 | 54 | 1992-1997 | Retrospective | Canada | The British Columbia Trauma Registry was searched and analyzed for patients incurring head injuries and length of hospital stay while skiing or snowboarding from January 1992 to December 1997 |
| Snowboarding/skiing | Skiing, Snowboarding, and Sledding Injuries in a Northwestern State | Carol S Federiuk, Jamie L Schlueter, Annette L Adams | Concussion Prevalence via Hospital/Urgent Care Records | Wilderness and Environmental Medicine | 2002 | 3 | 132 | 1992-1999 | Retrospective | USA | State trauma registry data from the 1992-93 through 1998-99 ski seasons on all snow sports participants transported to tertiary [illegible] |
| Soccer/Football | Concussions among university football and soccer players | J Scott Delaney, Vincent J Lacroix, Suzanne Leclerc, Karen M Johnston | Concussion Prevalence | Clinical Journal of Sports Med | 2002 | 4 | 529 | 1999-2000 | Retrospective survey | Cananda | A study to examine the incidence and characteristics of concussions among Canadian university athletes during 1 full year of football and soccer participation. |
| Softball | A Retrospective Analysis of Softball-Related Head and Facial Injuries Treated in United [illegible] | John S. Strickland, Marie Crandall, and Grant R. Bevill | Concussion Prevalence | Orthopedic Journal of Sports Med | 2019 | 3 | 3324 injuries | 2013-2017 | Descriptive Epidemiological Study | USA | To analyze head/face injury diagnoses and to identify the mechanisms associated with such [illegible] |
| Cricket | Situational factors associated with concussion in cricket identified from video analysis | Anna E Saw , David J Howard, Alex Kountouris, Andrew S McIntosh, John W Orchard, Richard Saw and Thomas Hill | Concussion Prevalence | Journal of Concussion | 2021 | 5 | 485 | 2015-2019 | retrospective case series analysis method | Australia | This study aimed to identify situational factors associated with concussion in elite Australian male and female cricket |
| Waterpolo | The Epidemiology of Sports-Related Head Injury and Concussion in Water Polo | Robert S. Blumenfeld, Jessica C. Winsell, James W. Hicks and Steven L. Small | Concussion Prevalence | Frontiers in Neurology | 2016 | 4 | 1470 | 2015 | Retrospective survey | USA | An electronic survey to the 44,000+ members of USA Water Polo, asking questions about concussions, head impacts, and symptoms commonly associ- ated with prior concussion. |
| Snowboarding/skiing | Emergency department reported head injuries from skiing and snowboarding among children and adolescents, 1996–2010 | Janessa M Graves, Jennifer M Whitehill, Joshua O Stream, Monica S Vavilala, Frederick P Rivara | Concussion Prevalence via Hospital/Urgent Care Records | BMJ Publishing Group | 2014 | 5 | 78538 | 1996- 2010 | Retrospective | USA | To evaluate the incidence of snow-sports-related head injuries among children and adolescents reported to emergency departments (EDs), and to examine the trend from 1996 to 2010 in ED visits for snow- |
| Snowboarding/Skiing | Concussion Among Youth Skiers and Snowboarders: A Review of the National Trauma Data Bank From 2009 to 2010 | Kelly R Bergmann, Andrew Flood, Nathaniel S Kreykes, Anupam B Kharbanda | Concussion Prevalence | Pediatric Emergency Care | 2015 | 4 | 1001 | 2009-2010 | Retrospective / cross sectional | USA | Subjects 18 years or younger with a ski- or snowboard-related injury were studied using data from the National Trauma Data Bank to determine concussion risk between helmet and non-helmet user |
| Baseball | Concussion rates and effects on player performance in Major League Baseball players | Vani J Sabesan, Beau Prey, Ryan Smith, Daniel J Lombardo,Wilfredo J Borroto, and James D Whaley | Concussion Prevalence | J Sports Med | 2018 | 5 | 112 | 2005-2016 | Descriptive Epidemiological Study | USA | Relative risk of concussion per 100,000 athlete exposures, independent t-test compare averages between groups. Paired sample t-tests were performed to compare pre- and postinjury statistics |
| Field Hockey | Concussion in field hockey: a retrospective analysis into the incidence rates, mechanisms, symptoms and recovery of concussive injuries sustained by elite field [illegible] | Michael Rossiter and Michael Challis | Concussion Prevalence | BMJ Open Sport and Exercise Science | 2017 | 2 | 28 | Not Reported | Retrospective | UK | To identify the incidence and mechanisms of concussion in elite Field Hockey in different age groups and also the post concussion symptoms and recovery times. |
| Hockey | Detailed description of Division I ice hockey concussions: Findings from the NCAA and Department of Defense CARE Consortium | Kathryn L. Van Pelt, Jaclyn B. Caccese, James T. Eckner, Margot Putukian, M. Alison Brooks, Kenneth L. Cameron, Megan N. Houston, Matthew A. Posner, Jonathan C. | Concussion Impact Factors | Journal of Sport Health Science | 2021 | 15 | 332 | 2014-2017 | Retrospective cohort study | USA | elucidate risk factors, specific mechanisms, and clinical presentations of concussion in men's and women's ice hockey. |
| Hockey | Concussion in Ice Hockey-A Cohort Study Across 29 Seasons | Mascha Pauelsen, Gusten Nyberg, Cecilia Tegner, Yelverton Tegner | Concussion Prevalence | Clinical Journal of Sports Med | 2017 | 4 | 267 | 1984-2013 | Cohort Study | Sweden | Incidence rate ratio for concussion and rehabilitation periods due to concussion were calculated and analyzed. |
| Skiing, Freestyle Skiing, snowboarding, motocross, dinghy sailing, sliding sports (luge, bobsleigh, skeleton), skating sports (figure, short track, speed) | The Incidence of Pediatric and Adolescent Concussion in Action Sports: A Systematic Review and Meta-Analysis | Francesco Feletti, and Matteo Bonato | Concussion Prevalence Systematic Review | Int J environmental ResPublic health | 2020 | 2 | 14 studies | 1980-2020 | Systematic/Meta | USA | This was a systematic review and meta-analysis of the incidence of concussion risk in youth athletes involved in action sports (AS |
| Soccer | Head and neck injuries in professional soccer | Michael Nilsson, Martin Hägglund, Jan Ekstrand, Markus Waldén | Concussion Prevalence | Clinical Journal of Sports Med | 2013 | 4 | 136 injuries | 2001-2010 | Prospective cohort | Sweden | Injury rate (number of time loss injuries per 1000 hours) |
| Luge | Injuries in the Sport of Luge: Epidemiology and Analysis | R S Cummings Jr 1 , A T Shurland, J A Prodoehl, K Moody, H H Sherk | Concussion Prevalence | The American Journal of Sports Medicine | 1997 | 5 | 1034 | 1985-1992 | Retrospective | USA | Between years 1985 -1992, data obtained from the athlete injury and illness report forms at the US Training Center Sports Medicine [illegible] |
| football, cheerleading, wrestling, volleyball, baseball, softball, boys' and girls' basketball, boys' and girls' soccer, and boys' and girls' track | Incidence and risk factors for concussion in high school athletes, North Carolina, 1996-1999 | Mark R Schulz, Stephen W Marshall, Frederick O Mueller, Jingzhen Yang, Nancy L Weaver, William D Kalsbeek, J Michael Bowling | Concussion Prevalence | American Journal of Epid | 2004 | 7 | 15802 | 1996-1990 | Prospective cohort | USA | Quantify risk factors for sports concussions |
| Baseball | Mild traumatic brain injury in major and Minor League Baseball players | Gary A. Green,Keshia M. Pollack, John D'Angelo,Mark S. Schickendantz, Roger Caplinger,, Kathleen Weber, Alex Valadka, Thomas W. McAllister, Randall W. Dick, Bert Mandelbaum,Frank C. Curriero, | Concussion Prevalence | American Journal of Sports Medicine | 2015 | 11 | 307 | 2011-2012 | Descriptive Epidemiology Study | USA | association between MTBI and return to play using several different measures. |
| Soccer | Mechanisms of head injuries in elite football | T E Andersen, A Arnason, L Engebretsen, R Bahr | Concussion Prevalence | British Journal of Sports Medicine | 2004 | 3 | 192 incidents | 1999-2000 | Prospective cohort | Norway | the aim of this study was to describe, using video analysis, the mechanisms of head injuries and of incidents with a high risk of head injury in elite football within Norway and Iceland. |
| Soccer | Concussion incidence and recovery in Swedish elite soccer — Prolonged recovery in female players | Fredrik Vedung, Sofie Hänni, Yelverton Tegner, Jakob Johansson, Niklas Marklund | Concussion Prevalence | Scandinavian Journal of Medicine & Science in Sports | 2020 | 5 | 959 | 2017 | Descriptive epidemiological study. | Sweden | In the 1st and 2nd soccer leagues for men and women, a Sport Concussion Assessment Tool (SCAT)-based questionnaire study was [illegible] |
| Soccer | Head injuries in the female football player: incidence, mechanisms, risk factors and management | Jiri Dvorak, Paul McCrory, Donald T Kirkendall | Concussion Prevalence | British Journal of Sports Med | 2007 | 2 | | 1998-2004 | Descriptive Epidemiological Study | USA | Exploring why females experience a higher rate of head injury, a different spectrum of injury and different mechanism of injury compared with male footballer |
| Soccer | Football injuries during the 2014 FIFA World Cup | Astrid Junge, Jiri Dvořák | Concussion Prevalence | British Journal of Sports Medicine | 2015 | 2 | 736 | 2014 | Retrospective | Switzerland | To analyse the incidence and characteristics of match injuries incurred during the 2014 FIFA World Cup in comparison to previous FIFA World Cups. |
| Soccer | Time Trends of Head Injuries Over Multiple Seasons in Professional Male Football (Soccer) | Florian Beaudouin, Karen Aus der Fünten, Tobias Tröß, Claus Reinsberger , Tim Meyer | Concussion Prevalence | Sports Med International Open | 2019 | 5 | 238 | 2006/07/ 2016-2017 | Prospective injury surveillance study | Germany | The present study aimed to investigate time trends of head injuries and their injury mechanisms since a rule change as monitoring may help to identify causes of head injuries and may advance head injury prevention efforts |
| Snowboarding/skiing | Head injuries among FIS World Cup alpine and freestyle skiers and snowboarders: a 7-year cohort study | Sophie E Steenstrup, Tone Bere, Roald Bahr | Concussion Prevalence | BJSM | 2013 | 3 | 2080 | 2006-2013 | Retrospective | Norway | retrospective interviews with FIS WC athletes at the end of seven consecutive seasons (2006-2013) to register injuries sustained during the competitive season to calculate the exposure of head and face injuries. |
| Cricket | Incidence of Concussion and Head Impacts in Australian Elite-Level Male and Female Cricketers After Head Impact Protocol Modifications | Thomas Hill, John Orchard, and Alex Kountouris | Concussion Prevalence | Sports Health | 2019 | 3 | 555 | 2015-2016 and 2016-2017 | Descriptive Epidemiological Study | Australia | subsequent analysis of head impacts and concussions in elite-level male and female cricketers in Australia over 2 seasons |
| CheerLeading | Catastrophic High School and Collegiate Cheerleading Injuries in the United States: An Examination of the 2006-2007 Basket Toss Rule Change | Rebecca K Yau , Savannah G Dennis, Barry P Boden, Robert C Cantu, James A Lord, Kristen L Kucera | Concussion Prevalence | Sports Health | 2019 | 6 | Basket tossing injuries n=19, subjects= 54 | 2002- 2017 | Case Series | USA | To describe catastrophic cheerleading injuries among high school and collegiate-level participants in the United States and to explore whether the 2006-2007 basket toss rule change was effective at reducing the number of catastrophic injuries |
| Football, soccer, volleyball, basketball, wrestling, baseball, softball | Concussions Among United States High School and Collegiate Athletes | Luke M Gessel, Sarah K Fields, Christy L Collins, Randall W Dick and R. Dawn Comstock | Concussion Prevalence | Journal of Athletic Training | 2007 | 5 | 4431 injuries | 2005-2006 | Descriptive Epidemiological Study | USA | To investigate the epidemiology of concussions in a nationally representative sample of high school athletes and to compare rates of concussion among high school and collegiate athletes. |
| Hockey | A prospective study of physician-observed concussions during junior ice hockey: implications for incidence rates | Paul Sean Echlin, Charles H Tator, Michael D Cusimano, Robert C Cantu, Jack E Taunton, Ross E G Upshur, Craig R Hall, Andrew M Johnson, Lorie A Forwell, Elaine N | Concussion Prevalence | Neurosurg Focus | 2010 | 10 | 67 | 2009-2010 | Prospective cohort | Canada | The objective of this study was to measure the incidence of concussion (scaled relative to number of athlete exposures) and recurrent concussion within 2 teams of fourth-tier junior ice hockey players (16-21 years old) during 1 |

| | | | | | | | | | | | |
|---|---|---|---|---|---|---|---|---|---|---|---|
| Volleyball | A Comparison of Women's Collegiate and Girls' High School Volleyball Injury Data Collected Prospectively Over a 4-Year Period | Jonathan C. Reeser, Andrew Gregory, Richard L. Berg,, R. Dawn Comstock, | Concussion Prevalence | American Ortho Society of Sports Med | 2015 | 4 | 729 High School Injuries/ 1380 College Injuries | 2005-2009 | Retrospective clinical review | USA | Analyze injury rates from collected data within the National Collegiate Athletic Association's Injury Surveillance System (NCAA ISS) and the High School Reporting Injuries Online (HS |
| Basketball | Epidemiology of Secondary School Boys' and Girls' Basketball Injuries: National Athletic Treatment, Injury and Outcomes Network | Alex N. Allen, Erin B. Wasserman, Richelle M. Williams, Janet E. Simon, Thomas P. Dompier, Zachary Y. Kerr, Alison R. Snyder Valier, PhD | Concussion Prevalence | Journal of Athletic Training | 2019 | 7 | 167 | 2011-2014 | Descriptive Epidemiological Study | USA | To describe the epidemiology of time-loss (TL) and NTL injuries sustained by secondary school boys' and girls' basketball athletes. |
| football, cheerleading, wrestling, diving/swim, baseball, softball, boys' and girls' basketball, boys' and girls' and boys' tennis and boys' and girls' track, golf, crew, girls soccer | Epidemiology of Sport-Related Concussions in High School Athletes: National Athletic Treatment, Injury and Outcomes Network (NATION), 2011–2012 Through 2013–2014 | Kathryn L. O'Connor, Melissa M. Baker, Sara L. Dalton, Thomas P. Dompier, Steven P. Broglio, and Zachary Y. Kerr | Concussion Prevalence | Journal of Athletic Training | 2017 | 6 | 2004 injuries | 2011-2014 | Descriptive Epidemiological Study | USA | To describe the epidemiology of sport-related concussion (SRC) in 27 high school sports during the 2011–2012 through 2013–2014 academic years. |
| Basketball | Effect of Mouthguard's on dental injuries and concussions in college basketball | Cynthia R Labella, Bryan W Smith, Asgeir Sigurdsson | Concussion Prevalence | Official Journal of the American College of Sports Medicine | 2001 | 3 | 70,936 | 1999-2000 | Survey | USA | During the 1999 to 2000 basketball season, athletic trainers from 50 men's Division I college basketball programs used an Internet |
| Football | Reported Concussion Rates for Three Division I Football Programs: An Evaluation of the New NCAA Concussion Policy | Kelly G. Kilcoyne, Jonathan F. Dickens, Steven J. Svoboda, Brett D. Owens, Kenneth L. CameronRobert T. Sullivan, John-Paul Rue | Concussion Prevalence | American Ortho Society of Sports Med | 2013 | 7 | 65 reported Concussions | 2009-2011 | Descriptive Epidemiological Study | USA | To determine the number of concussions that occurred on 3 collegiate Division I military academy football teams prior to and following recent changes in the NCAA concussion |
| boys' baseball, basketball, football, soccer, track, and wrestling; and girls' basketball, cheerleading, soccer, softball, track, and volleyball | Concussion Rates in U.S. Middle School Athletes, 2015–2016 School Year | Zachary Y .Kerr, Nelson Cortes, Amanda M.Caswell, Jatin P. Ambegaonkar, Kaitlin Romm Hallsmith, Frederick Milbert, Shane V.Caswell | Concussion Prevalence | American Journal of Sports med | 2017 | 7 | 2,622 | 2015-2016 | Descriptive Epidemiological Study | USA | Analyze concussion and athlete exposure (AE) data in school-sanctioned games and practices in boys' baseball, basketball, football, soccer, track, and wrestling; and girls' basketball, cheerleading, soccer, softball, track, and volleyball. |
| Gymnasstic/Cheer | Epidemiology of Cheerleading Stunt-Related Injuries in the United States | Brenda J Shields, Soledad A Fernandez, Gary A Smith | Concussion Prevalence | Journal of Athletic Training | 2009 | 3 | 338 | 2006-2007 | Prospective injury surveillance study | USA | The numbers, types, and rates of cheerleading stunt-related injuries during a 1-year period (2006-2007) are reported. Data retrieved from RIO (Reporting Information |
| Field Hockey | Head, Face, and Eye Injuries in Scholastic and Collegiate Lacrosse: A 4-Year Prospective Study | Andrew E. Lincoln, Richard Y. Hinton, Jon L. Almquist,Sean L. Lager, Randall W. Dick, | Concussion Prevalence | The American Journal of Sports Medicine | 2007 | 5 | 1,156,573 | 2000-2003 | Descriptive Epidemiological Study | USA | Injuries were analyzed in terms of (1) session type (practice or game), (2) nature of injury, (3) anatomical area injured, (4) mechanism of injury, (5) player activity at time of injury, and |
| Basketball | Epidemiology of Injuries in National Collegiate Athletic Association Men's Basketball: 2014–2015 Through 2018–2019 | Sarah N. Morris, Avinash Chandran, Landon B. Lempke, Adrian J. Boltz, Hannah J. Robison, Christy L. Collins | Concussion Prevalence | Journal of Athletic Training | 2021 | 6 | 3481 injuries total | 2014-2019 | Descriptive Epidemiological Study | USA | Injury counts and rates per 1000 AEs were examined by event type (practice, competition), competition level (Division I, |
| Football, soccer, volleyball, basketball, wrestling, baseball, softball | National High School Athlete Concussion Rates From 2005-2006 to 2011-2012 | Joseph A. Rosenthal,Randi E. Foraker, Christy L. Collins, R. Dawn Comstock | Concussion Prevalence | American Ortho Society of Sports Med | 2014 | 5 | 2516 Concussions reported | 2005-2012 | Descriptive Epidemiological Study | USA | During the academic years from 2005-2006 through 2011-2012, all injuries (including concussions) and exposure data from high school athletes participating in 9 sports were prospectively gathered by the HS RIO injury surveillance system, which has been |
| football, cheerleading, wrestling, volleyball, baseball, softball, boys' and girls' basketball, boys' and girls' soccer, and boys' and girls' track | Epidemiology of concussions among United States high school athletes in 20 sports | Mallika Marar, Natalie M McIlvain, Sarah K Fields, R Dawn Comstock | Concussion Prevalence | American Journal of Sports Medicine | 2012 | 4 | 14,635 | 2008-2010 | Retrospective cohort study | USA | In the 20 sports studied over the course of the 2008-2010 school years |
| Hockey | Incidence of Sports-Related Concussion Among NCAA Women's Ice Hockey Athletes | Emily M Brook, Emily Kroshus, Caroline H Hu, Marissa Gedman, Jamie E Collins, Elizabeth G Matzkin | Concussion Prevalence | Orthopedic Journal of Sports Medicine | 2017 | 6 | 459 | 2014-2015 | Descriptive Epidemiological Study | USA | To determine the incidence of sports-related concussion (SRC) in National Collegiate Athletic Association (NCAA) women's ice hockey athletes. |
| Hockey | Comparison of Concussion Rates Between NCAA Division I and Division III Men's and Women's Ice Hockey Players | John M Rosene, Bryan Raksnis, Brie Silva, Tyler Woefel, Paul S Visich, Thomas P Dompier, Zachary Y Kerr | Concussion Prevalence | American Journal of Sports Medicine | 2017 | 7 | 415 | 2009- 2015 | Descriptive Epidemiological Study | USA | To compare the epidemiologic patterns of concussion in National Collegiate Athletic Association (NCAA) ice hockey by sex and division. |
| Volleyball | Descriptive Epidemiology of Injuries Sustained in National Collegiate Athletic Association Men's and Women's Volleyball, 2013-2014 to 2014-2015 | Christine M Baugh, Gil S Weintraub, Andrew J Gregory, Aristarque Djoko, Thomas P Dompier, Zachary Y Kerr | Concussion Prevalence | Sports Health | 2018 | 6 | 18844 | 2014-2015 | Descriptive Epidemiological Study | USA | To examine injury epidemiology in NCAA men's and women's volleyball athletes. |
| Softball | Descriptive Epidemiology of Collegiate Women's Softball Injuries: National Collegiate Athletic Association Injury Surveillance System, 1988–1989 Through 2003–2004 | Stephen W Marshall, Karrie L Hamstra-Wright, Randall Dick, Katie A Grove, and Julie Agel | Concussion Prevalence | Journal of Athletic Training | 2007 | 5 | 5336 injuries | 1988-1989-2003-2004 | Descriptive Epidemiological Study | USA | The NCAA Injury Surveillance System has tracked injuries in all divisions of NCAA softbal |
| Basketball | Descriptive Epidemiology of Collegiate Women's Basketball Injuries: National Collegiate Athletic Association Injury Surveillance System, 1988–1989 Through 2003–2004 | Julie Agel, David E Olson, Randall Dick, Elizabeth A Arendt, Stephen W Marshall, and Robby S Sikka | Concussion Prevalence | Journal of Athletic Training | 2007 | 7 | 3556 Injuries | 1988-2004 | Retrospective | USA | To review 16 years of National Collegiate Athletic Association (NCAA) injury surveillance data for women's basketball and to identify potential areas for injury prevention initiatives. |
| Field Hockey | The First Decade of Web-Based Sports Injury Surveillance: Descriptive Epidemiology of Injuries in US High School Girls' Field Hockey (2008–2009 Through 2013–2014) and National Collegiate Athletic Association | Robert C. Lynall, Elizabeth C. Gardner, Jordan Paolucci,; Dustin W. Currie, Sarah B. Knowles, Lauren A. Pierpoint, Erin B. Wasserman, Thomas P. Dompier,. Dawn | Concussion Prevalence | The American Journal of Sports Medicine | 2015 | 1 | 150 | 2004-2009 | Descriptive Epidemiological Study | USA | To describe the epidemiology of injuries sustained in high school girls' field hockey in the 2008–2009 through 2013–2014 academic years and collegiate women's field hockey in the 2004–2005 through 2013–2014 academic |
| boys' football, wrestling, soccer, basketball, and baseball and girls' volleyball, soccer, basketball, and softball | Concussion Incidence and Trends in 20 High School Sports | Zachary Y. Kerr, Avinash Chandran, Aliza K. Nedimyer, Alan Arakkal, Lauren A. Pierpoint, Scott L. Zuckerman | Concussion Prevalence | American Academy of Pediatrics | 2019 | 6 | 6079 concussions | 2013-2014 & 2017-2018 | Descriptive Epidemiological Study | USA | Concussion rates per 10000 AEs with 95% confidence intervals (CIs) practice vs game<br><br>Linear regression |
| Football | Incidence of Concussion During Practice and Games in Youth, High School, and Collegiate American Football Players | Thomas P. Dompier, Zachary Y. Kerr, Stephen W. Marshall, Brian Hainline, Erin M. Snook, Ross Hayden, Janet | Concussion Prevalence | JAMA Pediatrics | 2015 | 7 | 20354 | 2012-2013 | Descriptive Epidemiological Study | USA | To examine and compare sport-related concussion outcomes (symptoms and return to play) in youth, high school, and collegiate football athletes. |
| Softball | Epidemiology of Injuries in National Collegiate Athletic Association Women's Softball: 2014–2015 Through 2018–2019 | Kevin L. Veillard, Adrian J. Boltz, Hannah J. Robison, Sarah N. Morris, Christy L. Collins, Avinash Chandran | Concussion Prevalence | Journal of Athletic Training | 2021 | 6 | 1511 injuries | 2014-2019 | Descriptive Epidemiological Study | USA | Exposure and injury data collected during competitive seasons in the NCAA Injury Surveillance Program during 2014–2015 through 2018–2019 (5 years) academic years |
| boys/girls—soccer and basketball, girls—field hockey and lacrosse, boys/girls—swim & dive, track & field, and | Concussion Incidence and Trends in 20 High | Zachary Y Kerr, Avinash Chandran, Aliza K Nedimyer, Alan Arakkal, Lauren A Pierpoint, Scott L Zuckerman | Concussion Prevalence | Pediatrics | 2019 | 6 | 9542 | 2013-2018 | Descriptive Epidemiological Study | USA | 2013-2014- 2017/2018 |

| Exposure Defined | Athletic Exposure Statistical Computation Reported | Statistical Analyses | Return to play time average (days) | Average concussions per athlete/ season | Incidence Rate Reporting | Stat Sig Reported |
|---|---|---|---|---|---|---|
| The injury rate was expressed per 1000 hours played | ∑ all injuries / ∑ all athlete-exposures | Regression Analysis of AE and Injury Rate | Not Reported | On average, there was a 6.1 concussion rate per 1000 hours | 6.5% BEACH and 7.5% INDOOR | P < 0.05 |
| Sum of concussive injuries per Season | - | T test on reported time loss following a concussion before and after implementation of concussion protocol | 5 games missed | 14.9 concussions per season | Not Reported | P < 0.05 |
| Sum of concussive injuries | Percentage of head injuries | Generalized Linear Model | Not Reported | In riders who suffered a fall from horseback that resulted in concussion (n=461), 33.4% contacted a hard floor surface such as asphalt, 26.5% landed on grass or dirt surface and 20.8% landed on indoor riding arena floor surfacing. Of these 461 riders, 88% reported they were wearing a helmet | 68.3% of head injuries occurred from horse handling, while mounting or dismounting only accounted for 1.5% | P < 0.05 |
| Sum of concussive injuries | Percentage of head injuries | Measures of central tendency were utilized to evaluate response patterns. | Over 51% (n = 19) took less than 5 days off, 30% (n = 11) took between 14 and 21 days and 8% (n = 3) took more than 30 days of | Not Reported | 45% sustained a concussion | Not Reported |
| Sum of concussive injuries | Percentage of head injuries | Multivariable regression analysis | Not reported | Not Reported | 45.2 TBI came from equestrian , 2.8% mortality rate in ICU | α = 0.05 yielding p < 0.01 |
| Sum of concussive injuries | Percentage of head injuries | Nationals concussion estimated risk by (%), year were quantified through spearman correlation coefficients | Not Reported | correlation of r = 0.97 . The total number of players experiencing a con-<br>cussion compared with the total number of players experi-<br>encing LoC over the 11-year time period had a correlation | National estimates were calculated to be 49,185 concus-<br>0 sions occurring in youth football players 5 to 13 years of age<br>over an 11-year time frame | P < 0.05, r > 0.618 |
| Sum of concussive injuries | Percentage of head injuries | Multivariate regression analysis | Eating disorder had an increased risk on concussion time-loss had an increased risk of time-loss injury (78% vs 65%; p=0.004) | Not Reported | 42% reported concussion history | P < 0.05 |
| Sum of concussive injuries | Percentage of head injuries | Multiple logistic regression was performed, and combined risk factors were calculated using inference trees. | Not reported | 127 reported concussions | 24% sustained a concussion | P <0.05 |
| Sum of concussive injuries | Percentage of head injuries | Systematic Review | Not Reported | Not Reported | Salt Lake City Olympics 2002- 19.6% of all patients (bobsled, 10.1%; luge, 8.7%; skeleton, 0.7%)<br><br>2010 Olympics (19), 0.07 of all registered athletes reported concussions, which was twice as high as the concussion incidence in the 2008 Summer Olympic Games.<br><br>2012 Youth Games - 0.07<br><br>At the 2014 Sochi Olympics (21), 11 concussions | Not Reported |
| Sum of concussive injuries | Percentage of head injuries | Descriptive Statistics | Both the hospital stay and intensive care unit stays were longer for the snowboarders with an average stay of 20.4 days on the ward and 11.8 days in the | Head injury was 0.005 per 1000 for skiers, 0.004 per 1000 for snowboarders, concussion was in 60% of skiers and 21% in snowboarders | M=.61 concussions per head injuries | Not Reported |
| Sum of concussive injuries | Percentage of head injuries | Regression Analysis of Odds Ratios | Not Reported | Concussions accounted for 39% of all injuries | Sledding concussions accounted for 55% of injuries while Skiing accounted for 39%, and snowboarding with the lowest reported percentage of 27% | Not Reported |
| Sum of concussive injuries | Percentage of head injuries | Logistic regression model using BIC | injuries leading to loss of time was 3.7 (95% CI 2.7 to 4.7) per 1000 player hours (men: 3.5, 95% CI 2.4 to 4.6; women: 4.1, 95% CI 2.1 to 6.1). | , 70.4% of the football players and 62.7% of the soccer players had experienced symptoms of a concussion during the previous year. Only 23.4% of the concussed football players and 19.8% of the concussed soccer players realized they had suffered a concussion. More than one concussion was experienced by 84.6% of the concussed football | head/neck injuries was 12.5 (95%CI 10.9 to 14.1) per 1000 player hours (men: 12.8, 95% CI 11.0 to 14.7; women: 11.5, 95% CI 8.4 to 14.6 | P < 0.05 |
| Sum of concussive injuries | Percentage of head injuries | Regression Analysis of head impacts | Not Reported | Concussions accounted for 17.7% of all injures | 84 reported female concussions and 16 reported male concussions | P <0.05 |
| risk of one head impact per every x amount of balls | Not Reported | Regression Analysis of head impacts | Median number of reported symptoms on the day of injury for concussed players was 7 out of 24 | Concussion was diagnosed in 35 cases (18%, 95% CI 13–24%) | Risk of one head impact every 7,736 balls or approximately 13 days play | P <0.002 |
| Percentage of head injuries | - | Regression Analysis of History of Concussion | Not Reported | Men reporting concussion sustained an average of 2.20 ± 0.12 concussions. Women reporting concussion sustained an average of 2.06 ± 0.08 concussions. | In this group of 534, respondents reported sustaining an average of 2.14, SE (±) = 0.07 concussions | P < 0.05 |
| per 'x' amount of resort visits | the number of head injuries divided by snow sports participation estimates derived from annual NSAA surveys | Regression Analysis of TBI visits | Not Reported | An estimated number of 78 538 (95% CI 66 350 to 90 727) snow sports-related head injuries among children and adolescents, 77.2% were traumatic brain injuries | The annual average incidence rate of TBI was 2.24 per 10 000 resort visits for children (4-12) compared with 3.13 per 10 000 visits for adolescents (13-17). | P < 0.05 |
| Odds Ratio | - | Multivariate regression analysis and odds ratio | Not Reported | Not Reported | Snowboarders had a greater likelihood of concussion compared to skiers (estimated-β, 2.1; 95% CI, 1.48-2.85) | P <0.05 |
| Not Reported | calculate the relative risk of concussion at the different positions, it was assumed that there were four catchers, eight infielders, eight outfielders, and 20 pitchers on each of the 30 team's rosters | Independent sample T-tests, paired sample | 31.3 days following concussions | Not Reported | Field players = 3 per 100,000 AE<br>Catchers 9 per 100,000 AE | P < 0.05 |
| Not Reported | ∑ all injuries / ∑ all athlete-exposures | Regression Analysis of AE | Not Reported | Not Reported | U16 boys- .63 per 1000hrs<br>U18 boys- .68 per 1000 hrs<br>U21 Men- 1.28 per 1000hrs<br>21 women - 3.03 per 1000hrs | Not Reported |
| Not Reported | Not Reported | Regression Analysis of Odds Ratios | Not Reported | Between 2014 and 2017, there were 332 athletes who participated in ice hockey; 47 sustained a concussion while enrolled in the CARE Consortium study | A prior concussion increased the odds of an incident concussion 2-fold (OR = 2.03; 95%CI: 1.04-4.08). | P <0.05 |
| Not Reported | Exposure = (Number of team-matches played * number of players per team* duration of match / 60) | Injury rate ratios (IRRs), χ2 statistics | Not Reported | 162 concussions | incidence rate ratio was 1.06 (confidence interval, 1.03-1.10) | P <0.05 |
| Not Reported | Not Reported | The incidence of concussion was reported in diverse ways, the two most common denominators being days of athletic exposure (DAEs) and player-year (PY) | Not Reported | alpine skiing (*n* = 10), freestyle skiing (cross, halfpipe, slopestyle, *n* = 2), snowboarding (halfpipe and slopestyle, *n* = 12), off-road motorcycle/motocross (*n* = 2), dinghy sailing (Europe and Laser classes, *n* = 1), sliding sports (bobsleigh, luge and skeleton, *n* = 2), skating sports (figure, short track and speed, *n* = 1 | overall incidence of concussion in children and adolescents aged ≤18 years involved in outdoor sports of 0.33/1000 DAEs | P < 0.05 |
| Not Reported | Not Reported | Simple and multiple risk factor analyses were evaluated using Cox regression for player-related | 10 days on average | head and neck injury during match play compared with training had a 78-fold higher rate of concussions from 2001 to 2010 | 0.06 concussions per 1000 AE hours. | IRRs with associated 95% CIs excluding 1.00 were considered statistically significant |
| Injuries per 'x' amount of runs | Not Reported | Injury Risk Ratio | The risk of sustaining any injury causing the loss of more than 1 day of practice was 0.04 per person per year, and 2% of all | 10 reported concussions out of 407 injuries | 2.5% sustained a concussion | Not Reported |
| If an athlete participated in any part of a game or practice, he or she was considered to have participated, and information was not collected on the degree of participation. | Not Reported | Generalized Poisson Regression Analysis of AE | Not Reported | overall rate of concussion was 17.15 (95% confidence interval: 13.30, 21.00) per 100,000 athlete-exposures | overall rate of concussion was 17.15 (95% confidence interval: 13.30, 21.00) per 100,000 athlete-exposures | IRRs with associated 95% CIs excluding 1.00 were considered statistically significant |
| Hours of game play | The average number of players per team per game on analysis of regular-season game participation via box scores that are publicly available. This average number over a season, multiplied by the number of team games at each professional level of baseball, was used as an estimate of game exposures | Regression Analysis of AE | 1w 2d | Not Reported | 0.42 per 1000 athletes | P < 0.05 |
| Hours of game play | Video recordings of incidents where a player appeared to be hit in the head and the match was consequently interrupted by the referee were analysed and cross referenced with reports of acute time loss injuries from the team medical staff | Injury Rate Ratios , Poisson regression | Not Reported | incidence of 1.7 per 1000 player hours (concussion incidence 0.5 per 1000 player hours) | (22.0 per 1000 player hours in Norway and 14.8 in Iceland, p = 0.009) | P < 0.05 |
| Hours of game play | Not Reported | Multivariate Regression | Not Reported | 1.19/1000 hours of game play | females 1.22/1000 hours vs males 1.18/1000 hours | P <0.05 |
| Hours of game play | Not Reported | Not Reported | Not Reported | Professional MALE<br>World Cup (6)- 51.0/1000<br>US (7)- 35.5 / 1000<br>Iceland (8)- 34.8/1000<br>UK (9) - 25.9/1000<br>Sweden (11)- 18/1000<br><br>Professional FEMALE<br>Sweden (12)- 24/1000<br>Germany (13/14)- 23.3/1000 | When concussions alone were considered, the incidence rate was 1.1/1000 player hours for men and 2.6/1000 player hours for women, | P < 0.05 95% CIs excluding 1.00 were considered statistically significant |
| Hours of game play | Match exposure was calculated by multiplying 1.5 h by 11 players and by the number of returned forms. | Reported % of injuries | Not Reported | Not Reported | Brazil(18%) - 5% concussed<br>S. Africa- 10%-1%, Germ 2006- 9%-1%<br>Japan 2002- 15%- 2%,<br>France 1998- 15%-1% | P < 0.05 |
| Hours of game play | Match exposure per team was calculated using the following calculation, number of games x number of players on the field (11 players) x duration of the game in hours (1.5h per match) | Regression Analysis of injury Rate Risk (IRR) | 06/07 - 5 (5)<br>07/08- 17 (16)<br>08/09- 12(10)<br>009/10- 8 (6)<br>10/11- 6(5)<br>11/12- 8(9)<br>12/13- 7(8)<br>13/14- 4(3)<br>14/15- 8(16)<br>15/16- 10(18)<br>16/17- 5(5)<br>TOTAL- 8(10) | 06/07- 0.41 [0.17–0.99]<br>07/08- 0.48 [0.22–1.07]<br>08/09- 0.32 [0.12–0.86]<br>009/10- 0.41 [0.17–0.98]<br>10/11- 0.58 [0.28–1.22]<br>11/12-0.49 [0.22–1.10]<br>12/13- 0.57 [0.27–1.19]<br>13/14-0.58 [0.28–1.22]<br>14/15- 0.49 [0.22–1.10<br>15/16- 0.81 [0.44–1.51]<br>16/17- 0.74 [0.39–1.42]<br>TOTAL- 0.53 [0.42–0.67] | Averaged time lost 10±10 days (median 6, range 1–47 days),<br>head injuries IR of 1.77 (95% CI 1.56–2.01) per 1000 match hours | P <0.05 |
| Head injuries were classified as 'head/face' injuries and did not include neck or cervical spine injuries. | To calculate the exposure, we extracted data from the official FIS website for all WC competitions for each of the athletes interviewed | Regression Analysis of Injury Rate Ratio (100 athletes per season) | Not Reported | 245 head/face injuries reported, nervous system injuries/concussions were the most common (81.6%) and 58 of these were severe (23.7%) | Highest in freestyle (5.7, 95% CI 4.5 to 6.8) and snowboard (5.0, 95% CI 4.0 to 6.0) compared with alpine skiing (3.5, 95% CI 2.7 to 4.4; RR 1.61, 95% CI 1.17 to 2.22 vs freestyle; RR 1.43, 95% CI 1.04 to 1.96 vs snowboard). | P <0.05 |
| head impacts/concussions per 1000 player days | Not Reported | Incidence calculation by player days | Not Reported | 92 head impacts that resulted in 29 concussions in all men's teams (on 1178 team days of play) and 15 head impacts that resulted in 8 concussions in women's matches (on 367 team days of play) | The match incidence rates were 7.2 head impacts (2.3 concussions) per 1000 player days in elite male cricket and 3.7 head impacts (2.0 concussions) per 1000 player days in elite female cricket | Not Reported |
| catastrophic injuries were those resulting from participating in skills related to the sport and were classified into categories of severity: serious (severe injuries without permanent functional disability), nonfatal (severe injuries with permanent functional disability), and fatal (injuries resulting in death) | Not Reported | Regression Analysis of catastrophic injuries | Not Reported | For basket tosses, injuries were most commonly sustained to the head (9/19; 47% | cheerleaders sustained serious injuries (n = 27; 50%) during practice (n = 37; 69%) to the head (n = 28; 52%) and cervical spine (n = 17; 32%). | P < 0.05 |
| Athlete-exposure (A-E) was defined as 1 athlete's participation in a practice or competition. | Not Reported | 1000 AE, Statistical analyses included calculation of rate ratios (RRs), proportion ratios (PRs), and χ2 tests | More than 50% of athletes in every sport returned to play in 9 days or less | Concussions accounted for 8.9% of all injuries | Girls HS - .25/1000<br>Girls College - 0.45/1000<br>Boys HS- .18/1000<br>Boys College- .38/1000 | All 95% confidence intervals (CIs) not containing 1.0 with P values of less than .05 were considered statistically significant. |
| Athlete exposures were defined within this sample as the sum of all games played, and were summed for all players. | Not Reported | The incidence of concussion was calculated as the number of observed or self-reported concussions occurring during observed games, divided by | Not Reported | incidence 21.5 concussions per 1000 athlete exposures) | A concussion was diagnosed by the physician in 19 (36.5%) of the 52 observed games | Not Reported |

| | | | | | | |
|---|---|---|---|---|---|---|
| athlete exposure (AE) as 1 athlete's participation in 1 competition or practice without regard for the duration of participation. | Not Reported | Regression Analysis of AE | The average severity of game-related concussion was similar for both the NCAA ISS and HS RIO: 88.1% of affected collegiate athletes were absent for 3 or | (NCAA ISS, 2.0 per 10,000 AEs; n = 69)<br>(HS RIO, 0.6 per 10,000 AEs; n = 38). | Concussions were 3.4 times more likely to occur in a collegiate volleyball player than in a high school player (injury rate ratio, 3.4; 95% CI, 2.3-5.1) | P < 0.05 |
| An *athlete-exposure* (AE) was defined as a single athlete participating in 1 secondary school-sanctioned practice or competition, regardless of duration, in which the athlete was exposed to the risk of injury. | ∑ all injuries / ∑ all athlete-exposures | Injury counts, rates, and rate ratios (IRRs) were reported with 95% confidence intervals (CIs). | Not Reported | Boys also experienced a lower concussion injury rate than girls for competitions (IRR = 0.48; 95% CI =0.33, 0.69) but not for practices (IRR = 0.77; 95% CI = 0.52, 1.16). | Boys vs girls in competition (IRR = 0.48; 95% CI =0.33, 0.69)<br>Boys vs girls in practices (IRR = 0.77; 95% CI = 0.52, 1.16). | All mean differences with 95% CIs not containing 0.0 were considered statistically significant. |
| An *athlete-exposure* (*AE*) was defined as 1 athlete participating in 1 practice or competition. Only those athletes with playing time in a competition were included in competition exposure calculations. | Not Reported | Regression model on RRs and IPRs were used to compare rates and proportions, respectively, in sex-comparable sports | Most athletes with SRCs reported symptom resolution within 7 days (40.7%, n = 814) or 8 to 14 days (21.7%, n = 23 | Reported Number of Concussions<br>Foot =1020,<br>Wrest = 137,<br>GField= 66,<br>GGYm= 8<br>GVoll= 74<br>Base= 18<br>Soft= 50<br>BBask= 92<br>Gbask= 128<br>Bcrew = 0<br>[illegible] | Overall rate of concussion was 3.89 per 10000 AEs among all sports | All 95% confidence intervals (CIs) not containing 1.0 with P values of less than .05 were considered statistically significant. |
| An athlete exposure was defined as one athlete participating in one practice or contest where he/she was exposed to the possibility of injury | Not Reported | Chi test Of difference in concussion exposure from mouth guard users to Monmouth guards | Not Reported | Not Reported | 0.52 per 1000 athletes | P < 0.05 |
| An athlete exposure was defined as 1 athlete participating in 1 practice or a game in which he was exposed to injury | ∑ all injuries / ∑ all athlete-exposures | Regression Analysis of Injury Rate Ratio | Not Reported | incidence rate across all 3 institutions was 2 times higher in 2010-2011 when compared with 2009-2010 (incidence rate ratio [IRR], 2.04; 95% CI, 1.20-3.55; P = 0.01) | Injury Risk 0.57 per 1000 athlete exposures in the 2009-2010<br>Injury Risk 1.16 per 1000 athlete exposures in the 2010-2011 | P <0.05 |
| An athlete exposure (AE) was defined as one athlete participating in one school-sanctioned game or practice. | Not Reported | Regression Analysis of AE and injury Rate Risk (IRR) | Not Reported | The overall concussion rate was higher in games than practices (1.15 vs 0.63/1,000 AEs, IRR=1.83, 95% CI=1.06, 3.15; | Game/Practice/Total/Game vs Practice 1000 AE's<br>Total Male- 1.12 (0.43, 1.81)<br><br>0.80 (0.48, 1.12)<br><br>0.87 (0.58, 1.17)<br><br>1.40 (0.67, 2.92)<br>Total Female- 1.18 (0.45, 1.91)<br><br>0.45 (0.20, 0.69) | All 95% confidence intervals (CIs) not containing 1.0. |
| An *AE* was defined as 1 cheerleader participating in 1 cheerleading event. | Not Reported | Multivariate logistic regression analysis's used, this involved the calculation of odds ratios (ORs) with 95% confidence intervals (CIs), χ2 | Not Reported | 6% of reported injuries were concussions | 0.04 injuries per 1000 AEs per practice and athletic events, 0.05 injuries per 1000 AEs | P ≤ .2 |
| An AE was defined as 1 athlete participating in 1 practice or game in which the high school or college participant was exposed to the possibility of injury | Not Reported | Regression Analysis of AE | Not Reported | Concussions constituted a higher percentage of injuries among boys (73%) and men (85%) than among girls (40%) and women (41%). | High School Female/Male ratio = 0.77<br>Collegiate Female/Male ratio = 0.86 | P < 0.05 |
| An AE was defined as 1 athlete participating in 1 exposure event. | Not Reported | Regression Analysis of AE and Injury Rate Ratios (IRR) | Not Reported | 3794 reported concussions over 5 years | 5.05% sustained a concussion | 95% CI excluding 1.00 |
| An AE was defined as 1 athlete participating in 1 competition or practice. | Not Reported | Regression Analysis of AE | Not Reported | CONCUSSIONS/1000 exposures<br>2006<br>Football- .47(.41, .53)<br>G.Soc- 0.36(.27,.47)<br>B.Soc- .22 (.16,.31)<br><br>[illegible] | By 2011-2012, the overall rate of concussions had increased significantly from 0.23 (0.21, 0.25) to 0.51 (0.48, 0.55) | <0.05 |
| An AE was 1 athlete participating in 1 athletic practice or competition. | Not Reported | Overall rates were calculated by dividing concussion incidence by AE using raw case counts.<br>rate ratios (RRs) and injury proportion ratios (IPRs) with P values and 95% confidence | 40% returned in 3 days or less, 2% returned the same day | 1289 concussions (66.6%) that occurred in competition and 647 (33.4%) that occurred during practice | concussion rate of 2.5 concussions per 10,000 AE. (RR, 5.7; 95% CI, 5.2-6.3)<br>girls - 1.6 per 10,000 AE<br>boys - 3.1 per 10,000 AE | IRRs with associated 95% CIs excluding 1.00 were considered statistically significant |
| An AE is defined as 1 athlete competing in 1 practice or game over a given period of time | It was assumed there were approximately 6 AEs per week over a 20-week season for NCAA Division II and III players and a 24-week season for Division I players and multiplied the number of athletes participating while considering their school's division | Regression Analysis of AE | Not Reported | (n = 219, 47.7%) of respondents reported at least 1 diagnosed concussion over the duration of their entire organized ice hockey career. A total of 13.3% (n = 61) of respondents reported a diagnosed concussion during the 2014-2015 season. | The incidence rate was 1.18 (95% CI, 0.92-1.51) per 1000 athlete-exposures to a game or practice and 0.58 (95% CI, 0.45-0.74) per 1000 hours of ice tim | P <0.05 |
| A reportable athlete-exposure (AE) was defined as 1 student-athlete participating in 1 NCAA-sanctioned practice or competition in which he or she was exposed to the possibility of athletic injury, regardless of the time associated with that participation. | Not Reported | Regression Analysis of AE and Injury Rate Ratios (IRR) and IPR | Not Reported | Division I men (0.83 per 1000 athlete-exposures [AEs]), followed by Division III women (0.78/1000 AEs), Division I women (0.65/1000 AEs), and Division III men (0.64/1000 AEs). | Division I men (0.83 per 1000 athlete-exposures [AEs]), Division III women (0.78/1000 AEs) | IRRs with associated 95% CIs excluding 1.00 were considered statistically significant |
| A reportable athlete-exposure (AE) was defined as 1 student-athlete participating in 1 NCAA-sanctioned practice or competition in which he or she was exposed to the possibility of athletic injury, regardless of the time associated with that participation | ∑ all injuries / ∑ all athlete-exposures | The injury rate was calculated as the number of injuries per 1000 AEs. Statistical analyses included calculation of injury rate ratios (IRRs | Not Reported | concussions (men, 19.4%; women, 14.8%) | 0.34 (0.07-0.61) men 1000 AE<br>0.39 (0.24-0.53) women | IRRs with associated 95% CIs excluding 1.00 were considered statistically significant |
| A reportable athlete-exposure (A-E) was defined as 1 student-athlete participating in 1 practice or competition in which he or she was exposed to the possibility of athletic injury, regardless of the time associated with that participation | ∑ all injuries / ∑ all athlete-exposures | Regression Analysis of AE | Not Reported | Concussions accounted for 6% of injuries | 0.25 game per 1000 AE<br>0.07 practice per 1000 AE | IRRs with associated 95% CIs excluding 1.00 were considered statistically significant |
| A reportable athlete-exposure (A-E) was defined as 1 student-athlete participating in 1 practice or competition in which he or she was exposed to the possibility of athletic injury, regardless of the time associated with that participation, Only participants with actual playing time were counted as having game exposures | ∑ all injuries / ∑ all athlete-exposures | Regression Analysis of AE | Not Reported | Games = (6.5% of all injuries) (95%CI, 0.43, 0.56)<br>Practices = (3.7%) of all injuries (95%CI, 0.13, 0.17) | Games - .50 per 1000 AE<br><br>Practices- 0.15 per 1000 AE | The 95% CIs provide information about the precision of each rate and can be used to determine if 2 rates differ statistically from one another. |
| A reportable *AE* was defined as 1 student-athlete participating in 1 school-sanctioned practice or competition in which he or she was exposed to the possibility of athletic injury, regardless of the time associated with that participation | Not Reported | Regression Analysis of AE and Injury Risk Ratios (IRR) | Nearly all of the players suffering a concussion were eventually able to return to play within the same season (92.8%). Of these, 77.3% returned in <10 days from the time of injury, while the other | Concussions accounted for 42.8% of all head, face, and eye injuries reported (0.40 per 1000 AEs; 95% CI, 0.32-0.53). | high school girls reported 1.73/1000 AEs (95% CI = 1.62, 1.83) vs college which reported 5.36/1000 AEs (95% CI = 5.02, 5.69) | All mean differences with 95% CIs not containing 0.0 were considered statistically significant. |
| A reportable AE was defined as 1 athlete participating in 1 school-sanctioned practice or competition. Our injury of focus, concussion, was defined as (1) occurring as a result of participation in an organized practice or competition, (2) requiring medical attention by an AT or physician, and (3) being diagnosed as a concussion. | Not Reported | Regression Analysis of Injury Proportion Rates and Injury Rate Risk and AE | Not Reported | AE by 10,000<br>Boys<br><br>Foot- 7.15 (6.73 to 7.61)<br><br>Ice - 16.24 (10.34 to 25.49<br><br>Lax- 8.96 (6.84 to 11.72) | 4.17 per 10000 AEs (95% CI: 4.09 to 4.26;) overall | IRRs with associated 95% CIs excluding 1.00 were considered statistically significant |
| 1 player participating in 1 game or practice | Not Reported | Descriptive analyses included the frequency and proportion of injuries. Comparative analyses included the calculation of risk (ie | Not Reported | most concussions at the high school and college levels occurred in practices (57.7% and 57.6%, respectively). In addition, no concussions were reported in youth football players aged 5 to 7 years, even though this age group contributed more than 7000 AEs | 9.6% youth, 4.0% HS, 8.0% in college sustained a concussion | CI 1.00 |
| 1 AE was defined as 1 athlete participating in 1 exposure event) | Not Reported | Injury rate ratios (IRRs) | Not Reported | 2223 reported concussions over 5 years | 5.5% sustained a concussion | IRRs with associated 95% CIs excluding 1.00 were considered statistically significant |
| Epidemiology of concussions in 20 high school sports during the 2013-2014 to 2017-2018 school years. | Not Reported | Regression Analysis of AE | - | overall rate of 4.17 per 10 000 AEs (95% CI: 4.09 to 4.26 | Boys' football had the highest overall concussion rate (10.40 per 10000 AEs), followed by girls' soccer (8.19 per 10000 AEs) and boys' ice hockey (7.69 per 10000 AEs | AE's with associated 95% CIs excluding 1.00 were considered statistically significant |

| Stat Significance Reporting Bt Exper Group/Control (p =) |
| --- |
| Indoor volleyball athletes had significantly higher injury rates compared with beach volleyball players for concussion (7.5% vs 6.5%; P < .0001) |
| 2006 to 2010 was 1.6, significantly less than the 5.0 games missed from 2011 to 2014, following the institution of the NBA concussion protocol (P = .023) |
| - The older the rider, the lower likelihood of suffering a head injury (OR=0.989, p<0.00005, 95% CI 0.985 to 0.993)<br>- Riders involved in only a single incident type had higher likelihood of suffering a head injury (OR=1.292, p=0.011, 95% CI 1.061 to 1.573). |
| Not Reported |
| Loss of consciousness was nonsignificant in equestrian sports ( p = .290)compared to roller blading, ski/snowboarding and aquatic based sports |
| Over an 11-year period there was a statistically significant increase r= 0.97. Additionally, The total number of players experiencing a concussion compared with the total number of players experiencing LoC over the 11-year time period had a correlation coefficient of r = 0.82 |
| Those with a concussion were more likely to have sought mental health treatment during college (32% vs. 23%; p=0.03), |
| Four factors impacted concussion risk<br>Older age (OR 1.03, 95% CI 1.01-1.06; p = 0.015)<br>Females (OR 2.54, 95% CI 1.04-6.21; p = 0.04)<br>having a diploma in horse riding vs not (OR 0.27, 95% CI 0.11-0.65; p = 0.004) |
| Not reported |
| Not reported |
| Skiers and snowboarders were less likely to have head injuries than sledders (odds ratio [OR] = 0.45; 95% CI, 0.21 to 0.98) |
| Soccer players were found to have an 11.1 times greater chance of suffering a concussion during the 1997 season if they had sustained a previous recognized concussion while playing soccer (p < 0.05). Football players who had suffered a previous recognized concussion not occurring during football were 4.2 times as likely to sustain a concussion (p < 0.05) |
| Not reported |
| Impact to the back of the head or helmet was the strongest factor suggestive of concussion (PPV 40%, p=0.028)<br><br>Not sig |
| Goalie status [$\beta = 0.58$, $t(639) = 3.829$, $p = 1.4 \times 10^{-4}$] and maximum level [$\beta = 0.41$, $t(639) = 8.25$, $p < 8.8 \times 10^{-16}$] significantly predicted the number of concussions |
| The incidence of TBI increased from 1996 to 2010 among adolescents (p<0.003). |
| Snowboarders had a greater likelihood of concussion compared to skiers (estimated-β, 2.1; 95% CI, 1.48-2.85) after adjusting for helmet status and age. Overall, Imputing missing values for helmets status had no effect on outcome for concussion. |
| The incidence rate of concussions increased significantly among players in all positions after the implementation of the 7-day DL rule in 2011 (P=0.003) |
| Not Reported |
| History of at least one prior concussion was significantly associated with incident concussion (Z = 2.03; p = 0.043) |
| It is shown that longer rehabilitation periods become more common over the years (P = 0.015) |
| Apart from motocross, sailing showed the highest incident rate per 1000 DAEs at 3.73 (95% CI: 0.31 to 7.15) |
| Risk Rate was found statistically significant (78.5; 95% CI, 24.4-252.5) |
| Not Reported |
| History of concussion was the only moderately strong risk factor for concussion (rate ratio = 2.28, 95% confidence interval: 1.24, 4.19). |
| p=0.42 |
| When comparing the ratios of the head injuries to head incidents between the two countries, Iceland had rates 2.15 times higher than Norway (p <0.0001). |
| No statistically significant difference in females vs males in incidence risk P = .85, but females vs males in return-to-play was statically significant (20 days vs 10 days, p = 0.02) |
| females were 1.57 times more likely to experience poor outcomes (eg, severe disability, persistent vegetative state) than males and more likely to die post- trauma (95% CI 1.09–2.82). |
| Not Reported |
| p=0.004. |
| Women had a higher injury incidence (5.8, 95% CI 4.8 to 6.9) versus men (3.9, 95% CI 3.2 to 4.6; RR 1.48, 95% CI 1.15 to 1.90) by 100 athletes |
| Not Reported |
| The overall basket toss injury rate (95% CI) was 0.75 (0.41-1.08) per 1,000,000 cheerleaders (0.76 [0.36-1.15] and 0.72 [0.09-1.36] for high school and college, respectively) |
| - Girls had a higher rate of concussion (0.36 concussions per 1000 A-Es) than boys (0.22 concussions per 1000 A-Es) (RR = 1.68, 95% CI = 1.08, 2.60, *P* = .03<br><br>- Girls had a higher rate of concussion (0.21 concussions per 1000 A-Es) than boys (0.07 concussions per 1000 A-Es) (RR = 2.93, 95% CI = 1.64, 5.24, *P* < .01).<br><br>- Concussions represented a significantly greater proportion of total injuries in softball players (5.5%, n = 3558) than in baseball players (2.9%, n = 1991) (PR = 1.91, 95% CI = 1.81, 2.01, *P* < .01 |
| Not Reported |

Table

| |
|---|
| NCAA ISS, 8.0% of the total injuries recorded [n = 111]; HS RIO, 3.8% of the total injuries recorded [n = 30]; P < 0.001. |
| No statistical difference between sexes |
| Concussion rate was higher in competition than in practice (RR = 3.30; 95% CI = 3.02, 3.60).<br><br>-Among sports in which both sexes participated (ie, baseball or softball, basketball, crew, cross-country, lacrosse, soccer, swimming and diving, tennis, indoor track and field, and outdoor track and field), the collective overall SRC rate was higher in girls than in boys (RR = 1.56; 95% CI = 1.34, 1.81)<br><br>- Symptom-resolution time greater than 14 days was higher in girls than in boys (33.0% [n = 125] versus 24.4% [n = 78]; IPR [illegible] |
| There were no significant differences between mouthguard users and nonusers in rates of concussions (0.35 vs 0.55) or oral soft tissue injuries (0.69 vs 1.06) |
| Significant difference from 2010-2011 vs 2009-2010 |
| - When considering sex-comparable sports only, the concussion rate was higher in girls than boys (0.66 vs 0.18/1,000 AEs, IRR=3.73, 95% CI=1.24, 11.23).<br><br>- Concussion rates did not vary between girls and boys (0.61 vs 0.87/1,000 AEs, IRR=0.70, 95% CI=0.41, 1.19 |
| collegiate teams were more likely to sustain a concussion or CHI than were cheerleaders on other types of teams (P = 0.02, OR = 3.10, 95% CI = 1.20, 8.06). |
| Sig in both collegiate and high school as both CI's crossed 1 |
| Not Reported |
| 5 of 9 showed a significant increase in concussion rates over time, including football (P = .002), boys basketball (P = .003), boys wrestling (P = .014), boys baseball (P = .036), and girls softball (P = .017) |
| girls had a higher concussion rate (1.7) than boys (1.0) (RR, 1.7; 95% CI, 1.4-2.0). |
| Diagnosed concussions over the 2014-2015 season were more likely to occur during the regular season (P < .001), in a game setting (P < .001), |
| only significant IRR was that the concussion rate was higher in Division I men than Division III men (IRR = 1.29; 95% CI, 1.02-1.65) |
| Not Reported |
| Concussion injury rate ratio was statically different in games vs practices (rate ratio = 3.6, 95% CI = 3.4, 3.8) |
| Concussion injury rate ratio was statically different in games vs practices (Rate Ratio= 3.3, 95% CI = 2.8, 4.0) |
| The injury rate from 2008–2009 through 2013–2014 was higher in college than in high school (3.25 versus 1.73/1000 AEs; IRR = 1.89; 95% CI = 1.63, 2.18). |
| the concussion rate was higher in competition than practice (10.37 vs 2.04 per 10000 AEs; IRR = 5.09; 95% CI: 4.89 to 5.31 |
| sole difference found was that the college concussion rate (0.53 per 1000 AEs) was lower than that in high school (0.66 per 1000 AEs; IRR, 0.80; 95% CI, 0.67-0.96) |
| Not Reported |
| among sex-comparable sports, girls had larger proportions of concussions that were recurrent than boys did (9.3% vs 6.4%; injury proportion ratio = 1.44; 95% CI: 1.11 to 1.88).<br>sex-comparable sports, concussion rates were higher in girls than in boys (3.35 vs 1.51 per 10,000 AEs; injury rate ratio = |
